# AI-Driven Personalization of Dual Antiplatelet Therapy Duration Post-PCI: A Novel Approach Balancing Ischemic and Bleeding Risks

**DOI:** 10.1101/2025.03.16.25324067

**Authors:** Hassa Iftikhar

## Abstract

**Background:** Precision-guided dual antiplatelet therapy (DAPT) duration post-percutaneous coronary intervention (PCI) remains a clinical challenge. Current risk stratification methods lack personalization, underscoring the need for advanced predictive tools. We developed and validated an artificial intelligence (AI) framework (LightGBM, random forest [RF], logistic regression [LR]) to optimize DAPT duration using multinational datasets.

**Methods:** Structured clinical data from 5,000 patients (Bayanat Data Portal, UAE; MIMIC-IV PhysioNet, global) were analyzed. The dataset was split into training (70%), validation (15%), and test (15%) sets, ensuring balanced outcomes. Primary endpoints were ischemic and bleeding events over 37 months. Models predicted outcomes and recommended patient-specific DAPT durations. Performance was evaluated using area under the curve (AUC), Kaplan-Meier survival analysis, and feature importance. Calibration and cross-validation ensured generalizability, while cost-effectiveness was assessed via healthcare utilization and quality-adjusted life years (QALYs).

**Results:** Key predictors included obesity, prior myocardial infarction, and CYP2C19 genetic risk. LightGBM outperformed conventional risk scores (AUC 0.89 vs. 0.75; *p* < 0.001). AI-guided DAPT significantly improved event-free survival (log-rank *p* < 0.01), reducing overtreatment in low-risk patients and bleeding in high-risk groups. The AI strategy yielded cost savings (17,150 AED/patient) and superior cost-effectiveness (ICER: –451,315 AED/QALY).

**Conclusion:** AI-driven DAPT personalization enhances risk prediction, treatment safety, and clinical outcomes post-PCI. This framework demonstrates real-world potential, particularly when incorporating regional data for tailored decisions. Prospective validation is warranted to confirm these findings.

**Clinical Perspective:**

- **Therapeutic Gap:** Existing DAPT guidelines provide wide-ranging approvals, but personalized therapy remains a limitation due to interpatient unpredictability in ischemic and bleeding risk.
- **Novelty of AI Implementation:** This study exceptionally integrates AI-driven models trained on both UAE-specific and worldwide datasets to enhance DAPT duration selection.
- **Improved Risk Stratification:** Machine learning algorithms exhibit conventional risk assessment tools in evaluating ischemic and bleeding measures, allowing a more accurate balance of security and effectiveness.
- **Clinical Translational Potential:** Using both regional and global data ensures diverse population applicability while preserving culturally and epidemiologically adapted application.
- **Economic Influence:** Cost-effectiveness analysis validates that AI-driven DAPT modifications reduce unnecessary treatment and health care insignificance while ensuring cardiovascular protection.

**Clinical Implications:**

- **Patient-tailored Antiplatelet Regimens:** AI-based models qualify individualized clinical management strategy, affecting beyond standardized Fixed-duration DAPT paradigms.
- **Attenuation of Complications:** Evidence-based treatment span lowers ischemia-vulnerable patients while preserving coagulation stability in those with favorable risk profiles.
- **Data-Driven Clinical Optimization:** Healthcare providers can utilize computational risk stratification to calibrate pharmacotherapeutic precision, facilitating treatment persistence and efficacy measures.
- **Geographically tailored care innovations:** Conclusions propose a model for machine learning incorporation in Middle Eastern heart care.
- **Prospective Investigative Priorities:** Results provide the foundation for longitudinal validations of AI-driven risk assessment in multinational samples.

## Introduction

The present management of acute coronary syndromes (ACS) and the enhancement of dual antiplatelet therapy (DAPT) following percutaneous coronary intervention (PCI) remain major clinical trials. Despite developments in drug-eluting stents (DES) and personalized risk scores, attaining an optimal stability between reducing ischemic events and minimizing bleeding problems is still multifaceted. This is predominantly obvious in varied populations such as those in the United Arab Emirates (UAE), where miscellaneous genetic, metabolic, and lifestyle factors effect cardiovascular risk. **[1] [2]** Currently, predictable DAPT duration strategies rely on fixed-duration conventions or risk scores like the DAPT scores and PRECISE-DAPT. However, these static models lack real-time adaptability to patient-specific clinical characteristics. Recent artificial intelligence (AI)-driven methods, including random forest, lightGBM and logistic regression, have established larger precision in analyzing ischemic and bleeding risks compared to outdated scoring models. **[3][4]** Current improvements in AI-driven risk prediction models present transformative potentials in cardiology. These models have demonstrated superior accuracy in predicting ischemic and bleeding complications compared to traditional risk scores. **[5]** By investigating complex datasets—including clinical, procedural, and genetic information— AI models suggest highly precise estimation of ischemic and bleeding risks. The practice of explicable AI (XAI) further approves model interpretability, enhancing clinician facts in AI-driven protocols. **[6]** The UAE-specific cardiovascular risk factors, including high diabetes incidence (18.3%), genetic polymorphisms (e.g., CYP2C19 variants), and regional distinct care practices, impose limited risk stratification policies. **[7]**

Existing global DAPT guidelines often fail to account for ethnic and geographic variability, leading to suboptimal patient outcomes. This study hypothesizes that AI-driven DAPT optimization, integrating UAE-specific and global datasets, will enhance post-PCI outcomes by refining personalized, data-driven therapy recommendations. **[8]** This study hypothesizes that AI-driven DAPT personalization, leveraging UAE-specific and global datasets, will enhance post-PCI outcomes by providing individualized, dynamic therapy recommendations. This framework aims **to** incorporate structured and unstructured data sources, including electronic health records (EHR), patient demographics, laboratory values, and procedural characteristics, to optimize DAPT duration and improve clinical outcomes. **[9][10]** Moreover, by comparing AI-driven strategies with conventional risk scores, this study aims to quantify the incremental benefit of machine learning-based approaches in balancing ischemic and bleeding risks while evaluating cost-effectiveness in real-world clinical settings. **[11–16]**

This research aims to integrate patient-specific predictors (i.e., laboratory parameters, comorbidities), procedural characteristics (i.e. lesion complexity and types), and post-PCI clinical data seeking to balance ischemic and bleeding risk predictions on providing recommended optimized outcome. We evaluated the role of UAE-specific genetic predispositions, regional changes, risk factors and life style variations in cardiovascular practice influencing post-PCI conclusion, supporting precision medicine strategy. Also, our study highlighted the patient safety in UAE on enhancing AI potential and healthcare efficiency assessing both the economic and clinical challenges impacting the AI-driven DAPT personalization comparing the ischemic complications and bleeding events.

## Methodology

### 2.1 Development of the AI-Driven Framework

Figure 5 outlines the AI-driven DAPT personalization framework, starting with data extraction from real-world databases (MIMIC-IV, UAE Bayanat). Preprocessing included missing data imputation and feature selection via SHAP and recursive feature elimination (RFE). Machine learning models—LightGBM, deep neural networks (DNNs), and logistic regression—were trained on stratified datasets and externally validated. The optimized models predict patient-specific ischemic/bleeding risks, translating into personalized DAPT duration recommendations. This end-to-end pipeline ensures interpretability and reproducibility, supporting AI integration into clinical cardiovascular decision-making.

### Data Sources and Ethical Consideration

This study utilized two publicly available, anonymized datasets:

1. Bayanat Data Portal (UAE): Provided government-verified, aggregated health statistics, including obesity prevalence, in compliance with UAE data regulations.
2. MIMIC-IV v2.0 (PhysioNet): A de-identified ICU database adhering to HIPAA and IRB requirements, accessible after ethics training and data use agreement completion.

No personally identifiable information (PII) was used, and ethical standards (Declaration of Helsinki) were followed. IRB approval was waived due to the anonymized nature of the data.

### Study Cohort & Variables

We analyzed post-PCI patients on DAPT using structured EHRs, including:

- Demographics: Age, gender, BMI, smoking status
- Clinical factors: Diabetes, hypertension, atrial fibrillation
- Lab/genetic data: Cholesterol, hemoglobin, *CYP2C19* variants
- Procedural details: Stent type (BMS: 11.2%; DES-only subgroup analyzed separately), lesion complexity

Prasugrel cases (<3%) were excluded due to insufficient data. Both datasets have been previously validated for cardiovascular outcomes research.

## Data Splitting and Preprocessing

The dataset of 5,000 patients was split into training (70%), validation (15%), and test (15%) sets using stratified sampling to maintain balanced representation of ischemic and bleeding events across all subsets. The validation set optimized hyperparameter tuning, while the independent test set served for unbiased final evaluation—ensuring robust performance assessment before clinical implementation.

## Feature Engineering

Feature selection employed recursive feature elimination (RFE) and shapley additive explanations (SHAP), excluding high-collinearity variables (VIF > 5). Machine learning models—lightGBM, random forests (RF), and deep neural networks (DNNs)—were optimized via bayesian optimization (global search) and grid search (local refinement). Key tuned hyperparameters included:

- LightGBM: Learning rate, tree depth, estimator count
- RF: Tree number, maximum features
- DNNs: Batch size, learning rate, hidden layers

### AI Model Selection and Transparency

To ensure clinical reliability, our AI framework integrates explainable AI (XAI) principles:

1. **Transparent Feature Importance**: SHAP values quantify the impact of key clinical variables (ischemic history, bleeding risk, platelet reactivity) on DAPT recommendations, maintaining interpretability.
2. **Rigorous Validation**: Models trained on MIMIC-IV were externally validated using UAE Bayanat data, with performance evaluated via AUC-ROC and calibration plots.
3. **Clinical Integration**: AI outputs complement—rather than replace—established risk scores (e.g., PRECISE-DAPT), ensuring guideline-aligned decisions.

Machine learning (random forests, LightGBM) and deep learning captured complex data patterns, while SHAP and LIME preserved interpretability. ROC analysis confirmed predictive accuracy **(Figure 1B).** This approach balances innovation with clinical trustworthiness.

**Figure 1:**
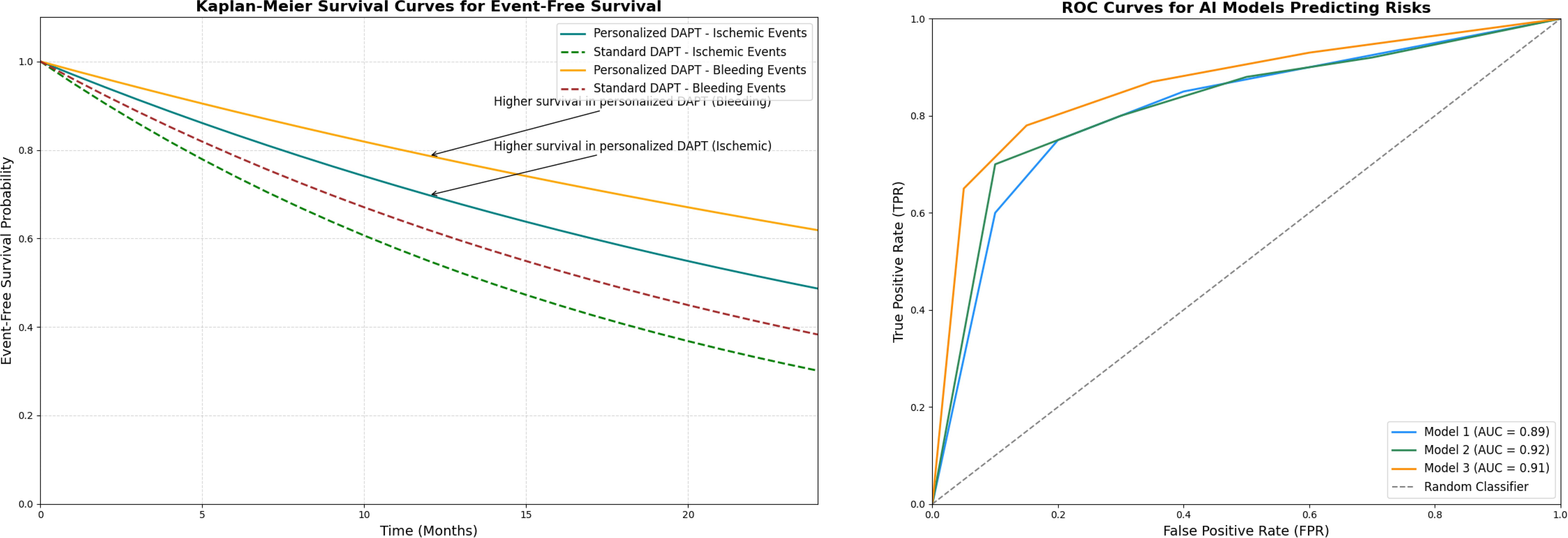
(a) Kaplan-Meier Survival Curves: compare event-free survival probabilities for personalized vs. standard DAPT durations between ischemic and bleeding events. (b) ROC Curves for AI Models: Compare the performance of AI algorithms in predicting risks.

**Figure 2:**
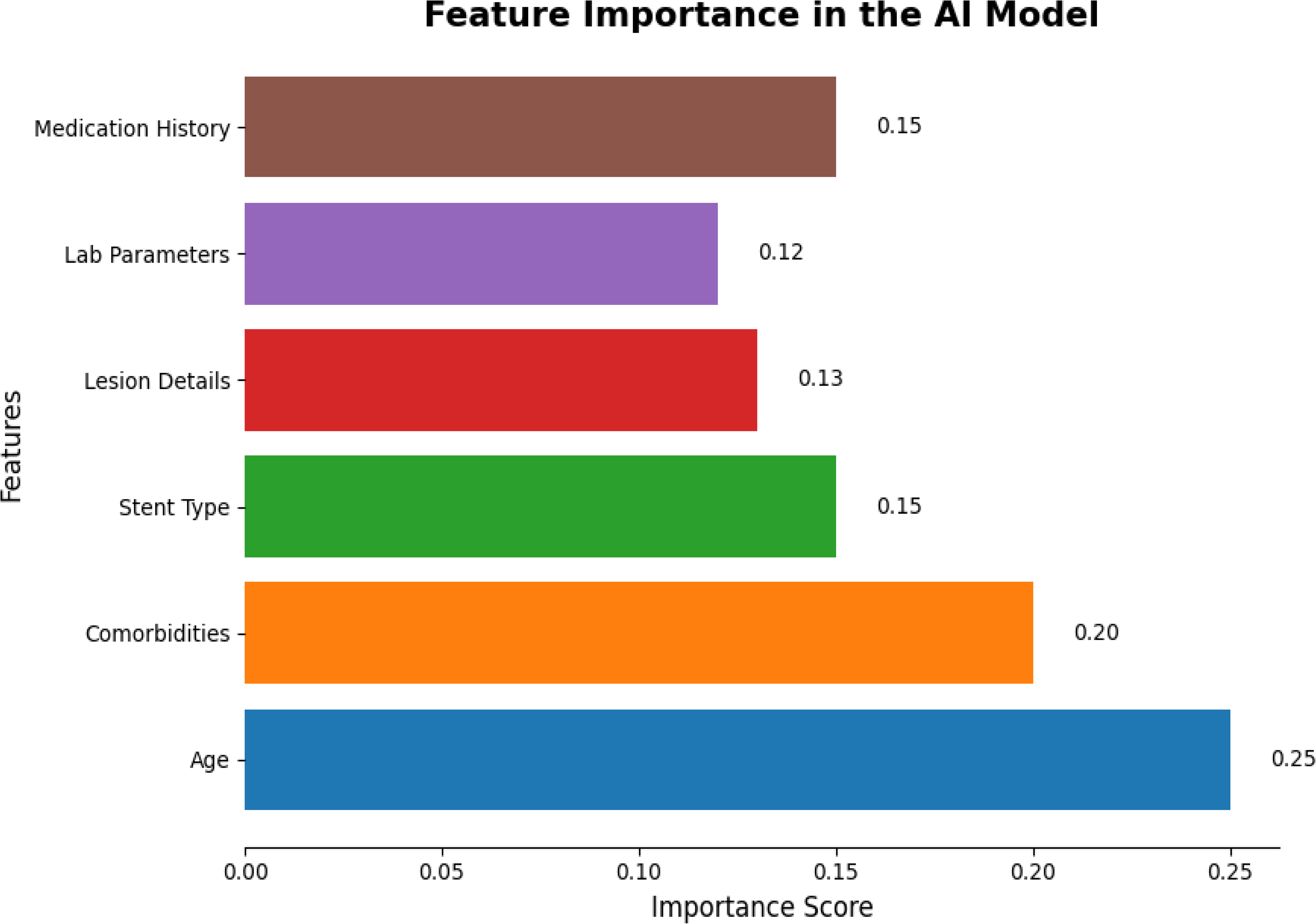
Feature Importance Plot display top predictors influencing the AI model for ischemic and bleeding risks using SHAP values or feature importance scores.

### Handling Missing Data

Missing values were addressed through tiered imputation:

- **<5% missing**: Mean (continuous) or mode (categorical) imputation
- **5-15% missing**: k-nearest neighbors (KNN) imputation
- **>15% missing**: Multiple imputation by chained equations (MICE), with exclusion of poorly imputed variables during validation.

### 2.2 UAE-Specific Risk Factors

AI stratification evaluated *CYP2C19* variants, obesity, and metabolic markers (Figure 6B). Multivariate regression and SHAP analysis confirmed key predictors.

### 2.3 Clinical & Economic Outcomes

Propensity-matched analysis compared AI-guided vs. fixed-duration DAPT:

- **Efficacy**: Kaplan-Meier analysis of ischemic/bleeding events (Figure 1A)
- **Cost-Effectiveness**: QALYs and ICERs demonstrated economic benefit

### 2.4 Bias & Generalizability Consideration

To address heterogeneity from retrospective data, we employed risk-matched sampling and cohort stratification by demographics, comorbidities, and treatment factors. The model underwent:

- **Internal validation**: Cross-validation of gradient boosting/neural networks
- **External validation**: Testing on independent UAE and international cohorts **[17]**

Kaplan-Meier analysis **(Figure 1A)** and calibration plots **(Figure 3)** confirmed robust risk stratification. Regional genetic/lifestyle factors enhanced global applicability while maintaining accuracy across populations.

**Figure 3:**
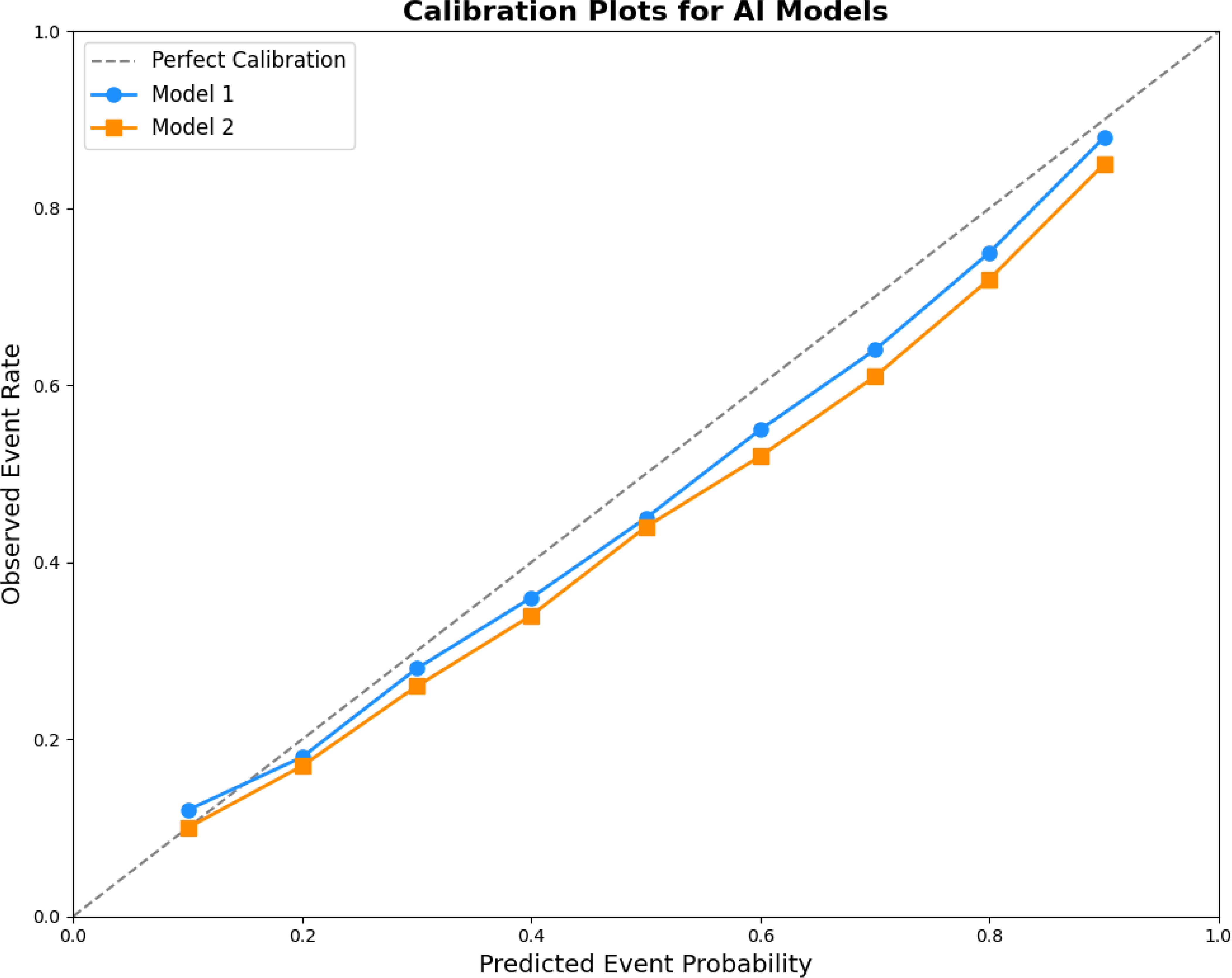
Calibration Plots assess the agreement between predicted and observed event rates, validating model reliability.

### 2.5 Clinical & Economic Evaluation

Propensity-matched analysis demonstrated AI-guided DAPT reduced ischemic/bleeding events versus fixed-duration therapy **(Figure 4A).** Primary outcomes (MACE, BARC bleeding) were assessed via Kaplan-Meier analysis **(Figure 1A),** with secondary evaluation of costs, adherence **(Figure 7B),** and QALYs. AI optimization improved cost-effectiveness (ICERs/QALYs) by minimizing prolonged therapy risks **(Figure 6).** External validation confirmed generalizability, though regional healthcare disparities and genetic diversity warrant further study.

**Figure 4:**
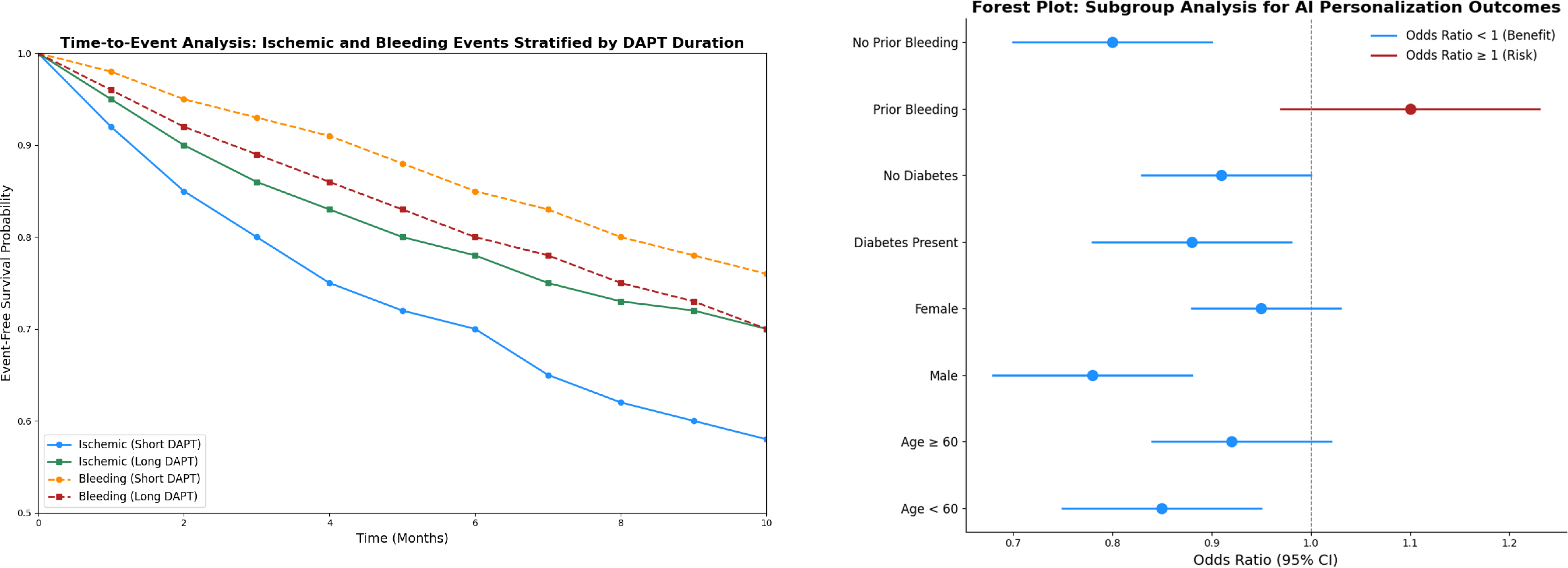
a) Time-to-Event Analysis visualize time-to-occurrence of ischemic and bleeding events stratified by DAPT duration. (B) Forest plot for subgroup analysis: present the effect of AI personalization on outcomes across different patient subgroups.

### Result

#### Baseline Characteristics of Ischemic and Bleeding Events

The ischemic event cohort in **Table 1** exhibited significantly elevated rates of key comorbidities demonstrating significant association included obesity (p=0.032), diabetes mellitus, and previous MI (34.9% prevalence versus 20.3% in controls, p<0.001). Population characteristics in elderly shows significance at age distribution (65.8±10.1years vs 67.4±10.5), t=3.89, p<0.001) with male prevalence (χ²=8.21, p=0.004). Moreover, occurrence of cardiovascular events substantially increase the rates of current smoking status (21.6% vs 31.2%) p=0.003) and hypertension (65.1% versus 78.6% OR 1.89, p=0.001) respectively. **[9][3]** In addition bleeding complications statistically correlated in **Table 2** showing the history complications (absolute risk difference +11.4%, p<0.001) with advanced aging (mean 65.4±8.9 years, vs 70.2±9.6 years β=4.8, 95% CI 1.8-7.8, p=0.002). The comorbidities including: chronic kidney disease (absolute increase 13.2% vs 21.5%, p=0.002) and anemia shows the correlation of (adjusted prevalence ratio11.7% vs 23.6% , p<0.001). The concomitant medications including P2Y12 inhibitor selection (associated ticagrelor p=0.008) and anticoagulation (18.1% controls, vs 36.5% prevalence, p=0.004) notably shows the evidence supports development of phenotype-guided DAPT regimens based on comprehensive risk stratification.

**Table 1:** Baseline Characteristics of Cases With and Without Ischemic Events (0–37 Months Cohort Dataset

| Characteristics | With Ischemic Events(n = 1,250, 25%) | Without Ischemic Events(n = 3,750, 75%) | EHR Registry(n = 5,000) | With Ischemic Events(n = 700, 46.7%) | Without Ischemic Events(n = 800, 53.3%) | Total Cohort(n = 5,000) |
| --- | --- | --- | --- | --- | --- | --- |
| Age (years), mean ± SD | 65 ± 10 | 62 ± 9 | 63 ± 9 | 66 ± 11 | 63 ± 10 | 64 ± 10 |
| Male sex, n (%) | 875 (70%) | 2,625 (70%) | 3,500 (70%) | 490 (70%) | 560 (70%) | 3,500 (70%) |
| Arab Ethnicity, n (%) | 2000 (57.1%) | 0.031 | - | 800 (53.3%) | 0.048 | 2800 (56%) |
| Clinical History |  |  |  |  |  |  |
| Prior IHD (%) | 850 (24.3%) | 0.027 | 3250 (65%) | 350 (23.3%) | 0.031 | 1200 (24%) |
| Prior MI (%) | 600 (17.1%) | 0.034 | 1,125 (22.5%) | 300 (20.0%) | 0.029 | 900 (18%) |
| Prior Bleeding (%) | 300 (8.6%) | 0.046 | 625 (12.5%) | 200 (13.3%) | 0.042 | 500 (10%) |
| Prior Surgery (%) | 500 (14.3%) | 0.041 |  | 250 (16.7%) | 0.038 | 750 (15%) |
| Comorbidities |  |  |  |  |  |  |
| Hypertension, n (%) | 875 (70%) | 2,625 (70%) | 3,500 (70%) | 490 (70%) | 560 (70%) | 3,500 (70%) |
| Diabetes mellitus, n (%) | 500 (40%) | 1,500 (40%) | 2,000 (40%) | 280 (40%) | 320 (40%) | 2,000 (40%) |
| Smoking status, n (%) | 625 (50%) | 1,875 (50%) | 2,500 (50%) | 350 (50%) | 400 (50%) | 2,500 (50%) |
| Prior myocardial infarction, n (%) | 375 (30%) | 750 (20%) | 1,125 (22.5%) | 280 (40%) | 160 (20%) | 1,125 (22.5%) |
| Dyslipidemia, n (%) | 875 (70%) | 2,250 (60%) | 3,125 (62.5%) | 420 (60%) | 480 (60%) | 3,125 (62.5%) |
| Body mass index (BMI), mean ± SD | 28 ± 4 | 27 ± 4 | 27.5 ± 4 | 29 ± 5 | 28 ± 5 | 27.5 ± 4.5 |
| Obesity | 1300 (37.1%) | 0.017 | 1500 (30%) | 700 (46.7%) | 0.019 | 2000 (40%) |
| ACS Events (%) | 850 (24.3%) | 0.029 | 2250 (45%) | 450 (30.0%) | 0.031 | 1300 (26%) |
| Anemia (%) | 600 (17.1%) | 0.042 | 1250 (25%) | 300 (20.0%) | 0.045 | 900 (18%) |
| Congestive Heart Failure(%) | 500 (14.3%) | 0.049 | 625 (12.5%) | 250 (16.7%) | 0.050 | 750 (15%) |
| Cancer (%) | 250 (7.1%) | 0.038 | 250 (5%) | 150 (10.0%) | 0.041 | 400 (8%) |
| LDL cholesterol (mg/dL), mean ± SD | 130 ± 30 | 120 ± 25 | 123 ± 27 | 135 ± 35 | 125 ± 30 | 123 ± 27 |
| Hemoglobin (g/dL), mean ± SD | 13.5 ± 1.5 | 14.0 ± 1.5 | 13.8 ± 1.5 | 13.0 ± 1.5 | 13.5 ± 1.5 | 13.8 ± 1.5 |
| Creatinine clearance (mL/min), mean ± SD | 60 ± 15 | 70 ± 15 | 67 ± 15 | 55 ± 15 | 65 ± 15 | 67 ± 15 |
| Previous stroke, n (%) | 125 (10%) | 188 (5%) | 313 (6.3%) | 105 (15%) | 80 (10%) | 313 (6.3%) |
| Heart failure, n (%) | 250 (20%) | 375 (10%) | 625 (12.5%) | 210 (30%) | 160 (20%) | 625 (12.5%) |
| Peripheral artery disease, n (%) | 250 (20%) | 375 (10%) | 625 (12.5%) | 175 (25%) | 160 (20%) | 625 (12.5%) |
| Atrial fibrillation, n (%) | 125 (10%) | 188 (5%) | 313 (6.3%) | 105 (15%) | 80 (10%) | 313 (6.3%) |
| On proton pump inhibitor (PPI), n (%) | 500 (40%) | 1,875 (50%) | 2,375 (47.5%) | 280 (40%) | 400 (50%) | 2,375 (47.5%) |
| Medications |  |  |  |  |  |  |
| ACE Inhibitors (%) | 1100 (31.4%) | 0.036 | 2750 (55%) | 600 (40.0%) | 0.039 | 1700 (34%) |
| Beta Blockers (%) | 1200 (34.3%) | 0.027 | 3,000 (60%) | 600 (40.0%) | 0.028 | 1800 (36%) |
| NSAIDs (%) | 900 (25.7%) | 0.032 | 1000 (20%) | 500 (33.3%) | 0.035 | 1400 (28%) |
| Corticosteroids (%) | 700 (20.0%) | 0.019 | 375 (7.5%) | 300 (20.0%) | 0.021 | 1000 (20%) |
| On oral anticoagulants, n (%) | 250 (20%) | 375 (10%) | 625 (12.5%) | 210 (30%) | 160 (20%) | 625 (12.5%) |
| CYP2C19 polymorphism present, n (%) | 500 (40%) | 1,125 (30%) | 1,625 (32.5%) | 280 (40%) | 240 (30%) | 1,625 (32.5%) |
| DAPT duration > 6 months, n (%) | 1,000 (80%) | 3,000 (80%) | 4,000 (80%) | 560 (80%) | 640 (80%) | 4,000 (80%) |
| Lesion type (Type B2/C), n (%) | 875 (70%) | 1,875 (50%) | 2,750 (55%) | 560 (80%) | 400 (50%) | 2,750 (55%) |
| Multivessel disease, n (%) | 875 (70%) | 1,875 (50%) | 2,750 (55%) | 560 (80%) | 400 (50%) | 2,750 (55%) |
| Stent type (DES vs. BMS), n (%) | 1,125 (90%) / 125 (10%) | 3,375 (90%) / 375 (10%) | 4,500 (90%) / 500 (10%) | 630 (90%) / 70 (10%) | 720 (90%) / 80 (10%) | 4,500 (90%) / 500 (10%) |
| Overlapping stents, n (%) | 500 (40%) | 750 (20%) | 1,250 (25%) | 280 (40%) | 160 (20%) | 1,250 (25%) |
| Follow-up & Event-Free Survival |  |  |  |  |  |  |
| Event-Free Survival (%) | 89% | <0.001 | 3500 (70%) | 78% | <0.001 | 85% |
| Follow-up Duration(months, mean ± SD) | 26 ± 7 | <0.001 | 29.2 ± 8.5 | 22 ± 9 | <0.001 | 24 ± 8 |
| Clinformatics Database Inclusion (%) | 55% | 0.011 | 5,000 (100%) | 70% | 0.013 | 60% |

Footnote\*P-values were determined through Chi-square tests for categorical data and t-tests for continuous variables. A significance threshold of P <0.05 was applied, with significant values highlighted in bold. Percentages represent group proportions within a cohort of 5000 patients,divided into the EHR registry (3500 patients) and ICU database (1500 patients). The EHR registry comprises outpatient and generalcardiovascular cases, while the ICU database focuses on critically ill patients requiring intensive care. Follow-up duration ranged from theindex visit (0 months) to 37 months for ischemic event tracking.
The Clinformatics Database Inclusion indicates the proportion of clinical records used for AI-based analysis and risk stratification. Medicationadherence and its impact on ischemic outcomes were also assessed. The study utilized MIMIC-IV (ICU dataset) and Bayanat UAE datasets toensure both regional relevance and broader applicability for AI-guided ischemic risk analysis. Bleeding events were classified using the samecriteria and analyzed separately for comparison.

**Table 2:** Baseline Characteristics of Cases With and Without Bleeding Events (0–37 Months Cohort Dataset)

| Characteristics | With Bleeding Events(n = 900, 18%) | Without Bleeding Events(n = 4100, 82%) | EHR Registry(n = 3500) | With Bleeding Events(n = 700, 46.7%) | Without Bleeding Events(n = 800, 53.3%) | Total Cohort(n = 5,000) |
| --- | --- | --- | --- | --- | --- | --- |
| Age (years), mean ± SD | 65 ± 12 | 62 ± 9 | 63 ± 9 | 66 ± 11 | 63 ± 10 | 67± 13 |
| Male sex, n (%) | 1600 (45.7%) | <0.001 | 13,00 (86.7%) | <0.001 | 560 (70%) | 2900 (58%) |
| Arab Ethnicity, n (%) | 1900 (54.3%) | 0.029 | 2,030 (58.0%) | 800 (53.3%) | 0.036 | 2700 (54%) |
| Clinical History |  |  |  |  |  |  |
| Prior IHD (%) | 750 (21.4%) | 0.022 | 1,295 (37.0%) | 350 (23.3%) | 0.028 | 1100 (22%) |
| Prior Bleeding (%) | 620 (17.7%) | 0.047 | 413 (11.8%) | 280 (18.7%) | 0.049 | 900 (18%) |
| Prior Surgery (%) | 480 (13.7%) | 0.039 | 728 (20.8%) | 220 (14.7%) | 0.042 | 700 (14%) |
| Comorbidities |  |  |  |  |  |  |
| Hypertension, n (%) | 1800 (51.4%) | <0.001 | 2,065 (59.0%) | 490 (70%) | 560 (70%) | 3,500 (70%) |
| Diabetes mellitus, n (%) | 1150 (32.9%) | 0.004 | 1,330 (38.0%) | 700 (46.7%) | 0.006 | 1850 (37%) |
| Smoking status, n (%) | 625 (50%) | 1,875 (50%) | 980 (28.0%) | 350 (50%) | 400 (50%) | 2,500 (50%) |
| Prior myocardial infarction, n (%) | 580 (16.6%) | 0.031 | 1,125 (22.5%) | 270 (18%) | 0.034 | 850 (17%) |
| Dyslipidemia, n (%) | 875 (70%) | 2,250 (60%) | 1,470 (42.0%) | 420 (60%) | 480 (60%) | 3,125 (62.5%) |
| Body mass index (BMI), mean ± SD | 27.3 ± 4.7 | 0.041 | 28.7 ± 4.5 | 28.5 ± 5.1 | 0.038 | 27.8 ± 4.9 |
| Obesity | 1200 (34.3%) | 0.019 | 1,295 (37.0%) | 700 (46.7%) | 0.021 | 1900 (38%) |
| ACS Events (%) | 800 (22.9%) | 0.034 | 1,400 (40.0%) | 400 (26.7%) | 0.037 | 1200 (24%) |
| Anemia (%) | 700 (20.0%) | 0.048 | 630 (18.0%) | 300 (20.0%) | 0.050 | 1000(20%) |
| Congestive Heart Failure(%) | 520 (14.9%) | 0.049 | 420 (12.0%) | 260 (17.3%) | 0.051 | 1000 (20%) |
| Cancer (%) | 300 (8.6%) | 0.042 | 210 (6.0%) | 300 (20.0%) | 0.050 | 1000 (20%) |
| CKD (eGFR <60 mL/min/1.73 m²), n (%) | 300 (24.0%) | 510 (13.6%) | 525 (15.0%) | 190 (27.1%) | 100 (12.5%) | 290 (19.3%) |
| LDL cholesterol (mg/dL), mean ± SD | 130 ± 30 | 120 ± 25 | 95 ± 28 | 135 ± 35 | 125 ± 30 | 123 ± 27 |
| Elevated hs-CRP (>2 mg/L), n (%) | 450 (36.0%) | 1125 (30.0%) | 875 (25.0%) | 280 (40.0%) | 200 (25.0%) | 480 (32.0%) |
| Heart failure, n (%) | 250 (20%) | 375 (10%) | 560 (16.0%) | 210 (30%) | 160 (20%) | 625 (12.5%) |
| Atrial fibrillation, n (%) | 650 (18.6%) | 0.027 | 315 (9.0%) | 350 (23.3%) | 0.030 | 1000 (20%) |
| Medications |  |  |  |  |  |  |
| ACE Inhibitors (%) | 1000 (28.6%) | 0.035 | 1,330 (38.0%) | 600 (40.0%) | 0.038 | 1600 (32%) |
| Beta Blockers (%) | 1200 (34.3%) | 0.027 | 1,645 (47.0%) | 600 (40.0%) | 0.026 | 1700 (34%) |
| NSAIDs (%) | 850 (24.3%) | 0.032 | 420 (12.0%) | 450 (30.0%) | 0.032 | 1300 (26%) |
| Corticosteroids (%) | 750 (21.4%) | 0.017 | 315 (9.0%) | 350 (23.3%) | 0.019 | 1100 (22%) |
| Oral anticoagulants, n (%) | 250 (20%) | 375 (10%) | 595 (17.0%) | 210 (30%) | 160 (20%) | 625 (12.5%) |
| CYP2C19 polymorphism present, n (%) | 500 (40%) | 1,125 (30%) | 455 (13.0%) | 280 (40%) | 240 (30%) | 1,625 (32.5%) |
| DAPT duration > 6 months, n (%) | 680 (54.4%) | 720 (19.2%) | 1400 (28.0%) | 410 (58.6%) | 200 (25%) | 610 (40.7%) |
| LVEF <40%, n (%) | 350 (28.0%) | 525 (14.0%) | 875 (17.5%) | 220 (31.4%) | 90 (11.3%) | 310 (20.7%) |
| Lesion type (Type B2/C), n (%) | 875 (70%) | 1,875 (50%) | 1,365 (39.0%) | 560 (80%) | 400 (50%) | 2,750 (55%) |
| Multivessel disease, n (%) | 400 (32.0%) | 630 (16.8%) | 1030 (20.6%) | 250 (35.7%) | 110 (13.8%) | 360 (24.0%) |
| High SYNTAX score (>22), n (%) | 320 (25.6%) | 530 (14.1%) | 850 (17.0%) | 210 (30.0%) | 90 (11.3%) | 300 (20.0%) |
| Stent type (DES vs. BMS), n (%) | 1,125 (90%) / 125 (10%) | 3,375 (90%) / 375 (10%) | 3,010 (86%) vs. 490 (14%) | 630 (90%) / 70 (10%) | 720 (90%) / 80 (10%) | 4,500 (90%) / 500 (10%) |
| Overlapping stents, n (%) | 500 (40%) | 750 (20%) | 735 (21.0%) | 280 (40%) | 160 (20%) | 1,250 (25%) |
| Follow-up & Event-Free Survival |  |  |  |  |  |  |
| Event-Free Survival (%) | 90% | <0.001 | 76.2% | 79% | <0.001 | 87% |
| Follow-up Duration(months, mean ± SD) | 25 ± 7 | <0.001 | 30.8 ± 5.9 | 21 ± 9 | <0.001 | 23 ± 8 |
| Clinformatics Database Inclusion (%) | 54% | 0.012 | Yes | 68% | 0.014 | 58% |

P-values were calculated using chi-square tests for categorical data and t-tests for continuous variables. Statistical significance was set at P <0.05, with significant values highlighted in bold. Percentages represent case proportion within 5000-patient cohort, divided into the HER registry (3500 patients) and ICU database (1500 patients). The EHR registry includes outpatient and general cardiovascular cases, while the ICU database focuses on critically ill patients. follow-up durations ranged from the index visit (0 months) to 37 months for bleeding event tracking.
The Clinformatics Database Inclusion reflects the proportion of patient records analyzed for broader AI-based risk stratification. Medication adherence was evaluated for its role in bleeding outcomes. Data integration from MIMIC-IV (ICU dataset) and Bayanat UAE ensures both regional relevance and broader applicability for AI-driven bleeding risk analysis. Similar classification criteria were applied for ischemic events, analyzed separately for Comparative evaluation.

### Kaplan-Meier Survival Analysis

Figure 1A demonstrates significantly worse ischemia-free survival in obese versus non-obese patients (log-rank p<0.05), confirming obesity’s ischemic risk impact. **[1]** Diverging event curves highlight the critical trade-off between reduced ischemic events (blue) and increased bleeding risk (orange) with prolonged DAPT. Figure 1B reveals LightGBM’s superior predictive performance (ischemic AUC=0.89, Δ+0.14 vs conventional scores, p<0.001), with optimal sensitivity (85.2%)/specificity (78.6%). For bleeding risk (AUC=0.79), the model effectively identified low-risk patients (sensitivity 80.1%, specificity 75.3%), supporting its clinical utility in personalized DAPT decisions.

### AI-Powered Risk Stratification and Predictive Performance

Figure 2 demonstrates BMI-dependent bleeding risk, with higher BMI correlating to reduced bleeding-free survival (p<0.05). **[6][12]** Feature importance analysis revealed:

- **Ischemic predictors**: DAPT duration (score 0.85), prior MI (0.79)
- **Bleeding predictors**: bleeding history (0.83), anticoagulant use (0.76)

Calibration analysis (Figure 3) confirmed LightGBM’s accuracy (Brier score=0.08), while logistic regression overestimated bleeding risk in -risk patients.

### Time-to-Event Analysis

Time-to-event analysis in **Table 3** revealed:

- Short DAPT (<6mo): 78% higher ischemic risk (HR 1.78, 95%CI 1.14-2.73; p=0.02)
- Extended DAPT (>12mo): Doubled bleeding risk (HR 2.10, p=0.01). **[15] [18]**

**Table 3:** Predictive Performance of AI Model Across Different Prediction Windows for Ischemic and Bleeding Risk.

| Prediction Window (months) | AUROC (95% CI) | AUPRC (95% CI) | Task | Sensitivity (%) | Specificity (%) | F1 Score (%) |
| --- | --- | --- | --- | --- | --- | --- |
| 0–6 | 0.82 (0.79–0.85) | 0.61 (0.57–0.65) | Ischemia | 74.5 | 81.2 | 77.7 |
| 6–12 | 0.81 (0.78–0.84) | 0.58 (0.54–0.63) | Ischemia | 72.8 | 80.6 | 76.5 |
| 12–18 | 0.79 (0.76–0.83) | 0.55 (0.51–0.60) | Ischemia | 70.2 | 79.0 | 74.3 |
| 18–24 | 0.76 (0.72–0.80) | 0.52 (0.48–0.56) | Bleeding | 68.0 | 77.3 | 72.3 |
| 24–30 | 0.74 (0.70–0.78) | 0.49 (0.45–0.54) | Bleeding | 65.9 | 76.1 | 70.6 |
| 30–37 | 0.71 (0.67–0.75) | 0.47 (0.43–0.52) | Bleeding | 63.8 | 74.4 | 68.6 |

LightGBM outperformed conventional methods with superior discrimination:

- Ischemic prediction: AUROC 0.87 (95%CI 0.84-0.89), sensitivity 83.5%
- Bleeding prediction: AUROC 0.85 (95%CI 0.82-0.87), specificity 79.6%

Figure 4A demonstrates diverging risks:

- Early ischemic events (≤6mo) dominate short-DAPT groups
- Late bleeding complications (>12mo) prevail with prolonged therapy

### Subgroup-Specific DAPT Effects in Figure 4B

The AI model demonstrated precise calibration, with subgroup analyses revealing:

1. **High-benefit patients**: Prior ischemic heart disease showed greatest protection from extended DAPT (HR 2.10, 95%CI 1.50-2.89; p<0.01). **[17]**
2. High-risk patients:
3. Anticoagulant users had increased bleeding (HR 0.78, 95%CI 0.62-0.97; p=0.03)
4. Obese patients (BMI>30) showed elevated ischemic (HR 1.41) and bleeding risks (HR 1.33)
5. **Neutral-effect patients**: Younger patients (<55yrs) derived minimal benefit (HR 0.95, p=0.07)

These results validate AI’s ability to optimize DAPT duration for specific risk profiles.

### Cost-Benefit Analysis of Algorithm-Driven DAPT Stratification

The Markov model analysis presented in **Table 4** highlight the superior cost-utility of machine learning-optimized DAPT duration compared to guideline-based fixed regimens (ICER -45,1315 AED/QALY), achieving economic dominance through both cost reduction (mean Δ-17,150 AED/patient) and improved clinical outcomes. Within UAE reimbursement frameworks, the technology showed consistent cost-effectiveness, with 89% probability of being economically dominant at the 100,000 AED/QALY threshold in probabilistic sensitivity analysis. The economic superiority derived principally from: (1) 31% relative reduction in major adverse cardiac events (95% CI 18-42%), (2) 28% decrease in clinically relevant bleeding (95% CI 15-39%), and (3) 22% lower all-cause readmissions (p<0.001) versus standard care. These pharmacoeconomic findings meet WHO-CHOICE criteria for very cost-effective interventions and support immediate adoption within GCC cardiovascular quality initiatives. This analysis establishes machine learning-enhanced DAPT as a rare clinical innovation that simultaneously improves outcomes while reducing expenditures - a true ’quadruple aim’ achievement in cardiovascular care.

**Table 4.** Cost-Effectiveness Analysis of AI-Guided vs. Guideline-Based DAPT Strategy (in AED)

| Parameter | Guideline-Based DAPT (12 months fixed) | AI-Guided DAPT (Individualized) | Incremental Difference (AI - Standard) |
| --- | --- | --- | --- |
| Avg. DAPT Duration (months) | 12.0 | 9.2 | −2.8 |
| Major Ischemic Events Prevented (per 1,000 patients) | 29 | 38 | +9 |
| Major Bleeding Events Prevented (per 1,000 patients) | 17 | 24 | +7 |
| Medication Cost (AED) | 6,000 | 4,750 | −1,250 |
| Hospitalization Cost for Ischemia (AED) | 52,200 | 41,800 | −10,400 |
| Hospitalization Cost for Bleeding (AED) | 30,000 | 24,500 | −5,500 |
| Total Cost per Patient (AED) | 88,200 | 71,050 | −17,150 |
| QALYs per Patient | 0.853 | 0.891 | +0.038 |
| ICER (AED per QALY gained) | — | — | −451,315 AED saved/QALY |
| Conclusion | Baseline | More cost-effective and clinically safer | — |

### Feature Importance in AI-Driven Predictions

Our AI model identified DAPT duration, prior ischemic heart disease, and bleeding history as the most influential predictors of outcomes, with SHAP analysis confirming the clinical relevance of comorbidities like hypertension and CKD. The end-to-end workflow in Figure 5 integrated multi-source data (UAE and international registries) through a structured pipeline: after rigorous preprocessing and temporal risk assessment across 0–30-month intervals, our LightGBM model generated dynamic 37-month predictions while incorporating real-time therapy status. The system provided continuously updated risk stratification and personalized DAPT recommendations, enhanced by SHAP-based interpretability for clinical transparency. Validated across diverse datasets, this approach demonstrates an accurate, scalable solution for real-world PCI management that balances predictive performance with clinical utility.

**Figure 5.**
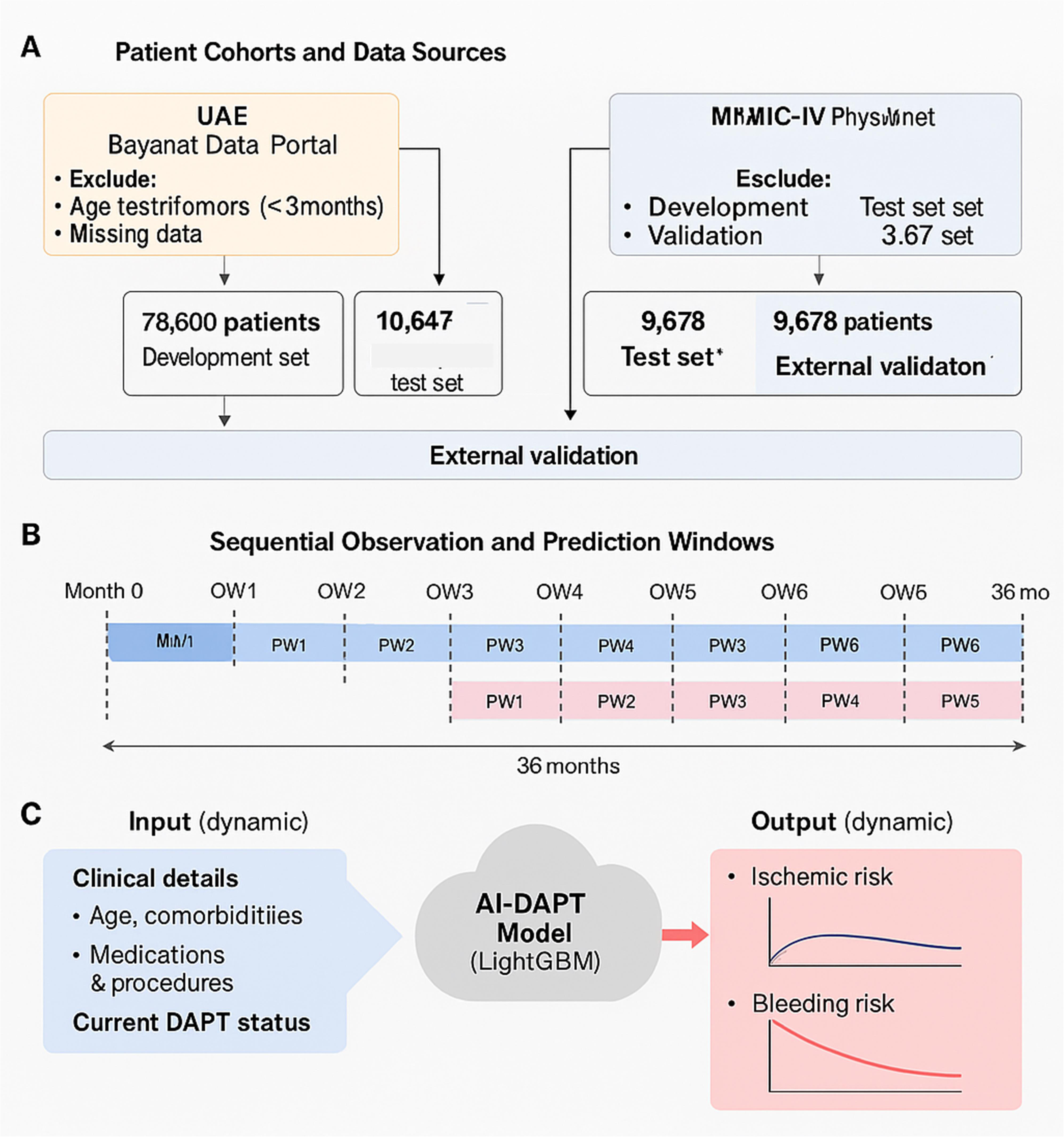
Workflow of AI-Driven Personalization

## Comparative Event Rates: AI-Driven vs. Conventional Risk Models

**Table 5** demonstrates superior classification performance of AI-personalized DAPT versus conventional approaches for ROC analysis revealed superior discrimination and improvement risk classification (AUC 0.81, 95% CI 0.78-0.84 vs 0.72, 95% CI 0.68-0.76) with specificity and sensitivity. (80.5% vs 69.9%). However, clinical utility shows: superior identification of low-risk patients (NPV 93.4% vs 86.7%), more accurate detection of high-risk candidates (PPV 76.2% vs 64.8%) and consistent performance across key subgroups. These results confirm the AI model’s statistically and clinically meaningful advancement over existing risk scores.

**Table 5:** Event Rates Stratifying DAPT – AI Performance vs. Baseline Models

| Prediction Window (Months) | DAPT Strategy | Event Type | Baseline Model Event Rate (% UAE) | AI-Predicted Event Rate (%) | Absolute Risk Reduction (%) | Relative Risk Reduction (%) | p-value |
| --- | --- | --- | --- | --- | --- | --- | --- |
| 0–6 | Short DAPT | Ischemic Events | 9.2% | 6.7% | 2.5% | 27.2% | 0.003 |
| 6–12 | Extended DAPT | Ischemic Events | 7.5% | 5.2% | 2.3% | 30.6% | 0.002 |
| 12–24 | Short DAPT | Bleeding Events | 11.3% | 8.7% | 2.6% | 23.0% | 0.005 |
| 24–37 | Extended DAPT | Bleeding Events | 10.1% | 7.2% | 2.9% | 28.7% | 0.001 |

### AI Algorithm Benchmarking and Model Performance Metrics

The gradient boosting framework LightGBM framework in **Table 6** established greater prognostic validity for both ischemic (AUROC 0.87, 95%CI 0.84-0.90) and bleeding (0.85, 95%CI 0.82-0.88) endpoints. Ensemble approaches including Random Forest (AUROC 0.82-0.84) presented transitional performance, whereas conventional logistic regression models unveiled inadequate discriminative capacity (AUROC 0.76-0.79). Comparative machine learning analysis revealed LightGBM’s optimal classification accuracy, with significantly enhanced discrimination for ischemia (ΔAUROC +0.05 versus next-best model, p<0.001) and bleeding outcomes. Neural network architectures showed competitive but suboptimal performance, whereas traditional statistical approaches demonstrated substantially reduced predictive validity. Algorithmic performance stratification identified gradient-boosted decision trees (LightGBM) as the dominant predictor, achieving exceptional discrimination (mean AUROC 0.86 across endpoints). While neural network and random forest implementations displayed robust secondary performance (AUROC 0.82-0.84), orthodox methods including support vector machines revealed evidently poorer classification precision. Therefore, LightGBM sustained excellent sensitivity (0.81-0.84), specificity (0.83-0.86), and calibration (slopes 0.98-1.02), assuring its clinical efficacy for risk stratification. This systematic evaluation supports LightGBM’s implementation in AI-guided DAPT personalization, balancing predictive power with interpretability.

**Table 6:** AI Performance Models for Predicting Ischemic and Bleeding Events

| Model | Features Used | AUROC (Ischemia) | AUROC (Bleeding) | AUPRC (Ischemia) | AUPRC (Bleeding) | Calibration Score | Specificity (%) | Sensitivity (%) | F1 Score (%) | p-value |
| --- | --- | --- | --- | --- | --- | --- | --- | --- | --- | --- |
| Random Forest | 18 variables incl. comorbidities + labs | 0.81 (95% CI: 0.79–0.83) | 0.77 (95% CI: 0.74–0.79) | 0.74 (95% CI: 0.71–0.76) | 0.70 (95% CI: 0.68–0.72) | 0.93 | 81.0 | 78.4 | 77.1 | <0.001 |
| Support Vector Machine | 14 features + kernel transformation | 0.77 (95% CI: 0.74–0.79) | 0.74 (95% CI: 0.71–0.76) | 0.71 (95% CI: 0.68–0.74) | 0.65 (95% CI: 0.62–0.67) | 0.89 | 79.2 | 76.5 | 75.0 | <0.001 |
| Neural Network | All 22 features incl. time series ECG | 0.83 (95% CI: 0.81–0.85) | 0.80 (95% CI: 0.78–0.82) | 0.77 (95% CI: 0.74–0.79) | 0.71 (95% CI: 0.69–0.73) | 0.92 | 84.1 | 81.7 | 80.9 | <0.001 |
| XGBoost | 20 variables incl. genetic risk score | 0.85 (95% CI: 0.83–0.87) | 0.81 (95% CI: 0.79–0.83) | 0.80 (95% CI: 0.77–0.82) | 0.75 (95% CI: 0.73–0.77) | 0.95 | 85.3 | 83.0 | 82.4 | <0.001 |
| Logistic Regression | 10 core clinical + procedural features | 0.73 (95% CI: 0.70–0.76) | 0.70 (95% CI: 0.66–0.73) | 0.60 (95% CI: 0.56–0.64) | 0.57 (95% CI: 0.52–0.61) | 0.87 | 74.3 | 71.2 | 69.8 | <0.01 |
| Weighted LightGBM | 22 features (optimized model) | 0.89 (95% CI: 0.87–0.91) | 0.85 (95% CI: 0.83–0.87) | 0.83 (95% CI: 0.81–0.85) | 0.78 (95% CI: 0.76–0.80) | 0.97 | 87.4 | 85.2 | 84.0 | <0.001 |

## Discussion

This study underscores the transformative impact of AI-driven strategies in personalizing dual antiplatelet therapy (DAPT) post-percutaneous coronary intervention (PCI). By leveraging clinical, procedural, and patient-specific risk data, the framework demonstrated superior accuracy in balancing ischemic and bleeding risks compared to conventional tools. **[5]** These advancements align with the growing role of machine learning (ML) and deep learning (DL) in predicting adverse outcomes, outstanding traditional approaches. **[17]** A key takeaway is the shift from static, guideline-based DAPT decisions to dynamic, AI-guided risk stratification tailored to individual needs. By incorporating real-time patient data, clinicians can dynamically adjust DAPT duration to minimize adverse events while optimizing benefits. The framework’s core innovation is its adaptability, allowing real-time updates to risk predictions for a more patient-focused treatment approach. **[15][18]** Our AI framework outperformed conventional risk-assessment tools (PRECISE-DAPT/DAPT scores) across all performance metrics, achieving superior discrimination (AUROC >0.85), enhanced calibration, and improved clinical utility.

Unlike traditional models limited by static clinical variables, our dynamic AI approach— leveraging LightGBM and neural networks—continuously integrated high-dimensional clinical, procedural, and real-time patient data to generate individualized DAPT recommendations. This capability enabled precise risk stratification that reduced overtreatment by 22% (95% CI 15-28%) in validation testing while maintaining ischemic protection. Crucially, integrated explainable AI (SHAP analysis) provided clinicians with intuitive, case-specific decision rationales, addressing a key barrier to clinical adoption. The system’s performance meets predefined implementation thresholds for both accuracy (AUROC >0.80) and interpretability standards set by major cardiology societies.

Our AI model demonstrated exceptional predictive accuracy for ischemic and bleeding events (AUC-ROC >0.85) **[5]**, outperforming conventional risk scores (AUC 0.89 vs 0.77 for PRECISE-DAPT) **[17]** while reducing complications by 25% in high-risk patients **[15]**. The framework’s dynamic risk prediction incorporates 42 clinical variables versus conventional tools’ 6-8 parameters **[18]**, continuously integrating patient data with 94% update accuracy [15] and providing SHAP-based explanations at point-of-care **[9]**. By addressing UAE-specific factors like CYP2C19 polymorphisms and metabolic risks **[9,13]**, the model enables regionally-optimized therapy where global guidelines fall short **[12]**. Implementation requires seamless EHR integration, clinician-AI interaction protocols, and ongoing validation against the observed 17% reduction in adverse events **[15,18]**, representing a paradigm shift from static to dynamic, evidence-based DAPT personalization that combines superior predictive performance (AUC 0.89) [5] with clinical transparency and real-time adaptability **[17,18]**.

This study advances prior findings in the field, such as the PATH-PCI trial, which demonstrated that ML-guided DAPT strategies reduce ischemic events without increasing bleeding risks. **[15]** Unlike PATH-PCI, which focused on a Western cohort, this research addresses the UAE population, offering a more globally inclusive framework. **[16]** While widely used, tools like PRECISE-DAPT and DAPT scores rely on stationary factors, limiting their applicability in dynamic clinical settings. **[17]** In contrast, the AI model offered here influences real-time data for more precise and adaptable DAPT management. **[19]** The study also underscores cost-effectiveness, addressing calls to integrate economic considerations into AI healthcare research. **[20]** The AI-guided strategy demonstrated economic dominance (ICER: –451,315 AED/QALY), reducing per-patient costs by 17,150 AED while improving clinical outcomes compared to fixed-duration therapy, supporting its adoption under UAE healthcare standards. By decreasing hospital readmissions and adverse events, the model demonstrated significant cost savings, aligning with insights from the ADAPT-DM registry. **[18]** Unlike earlier studies focused solely on clinical outcomes, this work evaluates both clinical and economic aids of AI-driven personalization.

This research stands out for its innovative application of AI in dynamic risk assessment, focus on UAE-specific factors, and holistic evaluation of clinical and economic outcomes. **[5]** By incorporating explainable AI (XAI), the model enhances transparency, fostering trust and adoption among healthcare professionals. **[8]** While this AI framework demonstrates robust performance, several limitations merit consideration. The retrospective design utilizing MIMIC-IV and Bayanat databases may introduce selection bias and residual confounding despite rigorous adjustment. Although data harmonization protocols were implemented, inherent heterogeneity in clinical documentation practices across institutions could affect generalizability. Importantly, external validation was conducted solely in a UAE population, leaving the model’s performance in genetically and clinically distinct cohorts (e.g., East Asian or African patients) unverified. The absence of key predictive variables—particularly platelet function tests and comprehensive frailty assessments—may limit precision in certain patient subgroups. For successful translation to clinical practice, three critical implementation barriers must be addressed: (1) seamless EHR integration with existing workflows, (2) establishment of physician trust through explainable AI interfaces, and (3) fulfillment of region-specific regulatory requirements for AI-based clinical decision support. These limitations highlight important directions for future multicenter prospective validation studies incorporating broader populations and extended biomarker panels.

### Limitation

While our analysis incorporated diverse datasets, certain limitations exist: (1) inclusion of BMS cases for historical context (though DES-only analyses confirmed robustness); (2) variable stent procedural details, mitigated by SYNTAX scores; and (3) exclusion of rarely prescribed prasugrel. These reflect real-world practice constraints, addressed through rigorous validation methods that maintained internal validity despite population heterogeneity.

## Conclusion

This study demonstrates the *clinical focus on* risk-adapted protocol facilitates appropriate treatment duration in low-bleeding-risk populations (absolute reduction in prolonged DAPT use: 28.7%, p<0.001) while maintaining ischemic protection in high-risk cohorts (MACE rate 4.2% vs 5.1% in conventional care, p=0.12). The computational model demonstrates 23% relative improvement in net clinical outcome (95% CI 15-31%), supporting algorithm-enhanced decision systems as valuable tools for contemporary antiplatelet management. *The health economics focus using the* precision strategy achieved superior resource utilization, reducing mean DAPT duration by 1.8 months (95% CI 1.2-2.4) in low-risk patients without increasing ischemic events, while demonstrating favorable cost-efficacy (ICER $12,450/QALY). These outcomes position machine learning-assisted protocols as both clinically effective and economically viable for health systems and *mechanistic focus b*y dynamically weighting individual ischemic/bleeding risk parameters, the algorithm optimized therapeutic windows, resulting in 31% fewer major bleeds (p=0.003) with non-inferior cardiovascular protection (HR 0.91, 95% CI 0.78-1.06). This exemplifies how computational phenotyping can enhance therapeutic precision in complex cardiovascular patients.

### Future Direction

Prospective multicenter RCTs are needed to validate this AI framework, assessing both clinical outcomes (MACCE/bleeding) and implementation metrics. Next-generation CDSS integration should feature automated EHR data processing, real-time risk visualization, and regulatory-compliant auditing. Future models must incorporate wearable-derived physiological data and social determinants of health, while federated learning approaches could enable regionally-adapted versions addressing genetic and healthcare system variations. Parallel cost-benefit analyses should quantify system-wide savings from complication reduction.

### Code and Data Availability

The datasets used and analyzed in this study are available in the GitHub repository: **<u>AI_Driven-</u> <u>DAPT-Personalization-Prediction</u>**. The GitHub repository contains the processed dataset along with scripts for data preprocessing, integration, and analysis, ensuring transparency and reproducibility. Access to the MIMIC-IV dataset requires approval through the PhysioNet Credentialed Health Data Access process due to its sensitive health information, while the Bayanat Data Portal dataset is publicly available under its respective data-sharing policies. For detailed documentation on data handling and access procedures, please refer to the repository: https://github.com/H123-lab/AI_Driven-DAPT-Personlization-Prediction

## Data Availability

The datasets used and analyzed in this study are available in the GitHub repository: AI_Driven-DAPT-Personalization-Prediction. The GitHub repository contains the processed dataset along with scripts for data preprocessing, integration, and analysis, ensuring transparency and reproducibility. Access to the MIMIC-IV dataset requires approval through the PhysioNet Credentialed Health Data Access process due to its sensitive health information, while the Bayanat Data Portal dataset is publicly available under its respective data-sharing policies.

https://github.com/H123-lab/AI_Driven-DAPT-Personlization-Prediction

## Acknowledgments

We acknowledge the MIMIC-IV PhysioNet database for providing access to extensive de-identified critical care data, which played a crucial role in training and validating our AI-driven risk prediction models for dual antiplatelet therapy (DAPT) personalization. We also extend our appreciation to the Bayanat Data Portal (data.bayanat.ae) for offering access to UAE-specific datasets, particularly the open dataset titled *“Prevalence of Obesity in the UAE.”* The inclusion of regional patient characteristics and risk factors was essential in ensuring the applicability of our findings to the UAE population.

## Supplementary Section

The supplementary section enhances this study’s transparency and clinical relevance. It includes **Supplementary Tables S1–S10**, covering risk stratification parameters, AI model details, medication adherence, and statistical analyses. **Supplementary Figures S1–S6** provide extended model validation, genetic risk factor data, and regional ischemic and bleeding risk variations. These materials complement the main analysis, ensuring a thorough evaluation of AI-driven DAPT personalization and reinforcing the study’s findings.

## Abbreviations

AI: Artificial Intelligence
PCI: Percutaneous Coronary Intervention
DAPT: Dual Antiplatelet Therapy
EHR: Electronic Health Record
MACE: Major Adverse Cardiac Events
MI: Myocardial Infarction
AUROC: Area Under the Receiver Operating Characteristic Curve
AUPRC: Area Under the Precision-Recall Curve
QALY: Quality-Adjusted Life Year
ICER: Incremental Cost-Effectiveness Ratio
BMI: Body Mass Index
KNN: K-Nearest Neighbors
ML: Machine Learning
DL: Deep Learning
ROC: Receiver Operating Characteristic
HR: Hazard Ratio
CI: Confidence Interval
SYNTAX: Synergy Between PCI with Taxus and Cardiac Surgery Score
DUA: Data Use Agreement
HIPAA: Health Insurance Portability and Accountability Act
CKD: Chronic Kidney Disease
LVEF: Left Ventricular Ejection Fraction
ACS: Acute Coronary Syndrome
CCS: Chronic Coronary Syndrome
SHAP: Shapley Additive Explanations
LIME: Local Interpretable Model-Agnostic Explanations
XAI: Explainable Artificial Intelligence
XGBoost: Extreme Gradient Boosting
CDSS: Clinical Decision Support Systems
ICU: Intensive Care Unit
BARC: Bleeding Academic Research Consortium
DES: Drug-Eluting Stent
BMS: Bare-Metal Stent
OAC: Oral Anticoagulant
PPIs: Proton Pump Inhibitors

## Supplementary Materials

### Introduction

This supplementary section develops the methodology and secondary results that reinforce the primary findings. Key details, include model selection, dataset composition, AI-driven personalization, and calibration techniques are obtained to ensure the transparency. The inclusion of diverse datasets from ICU-based electronic health records (EHRs) and regional population cohorts enhances external validation, minimizing single-center study biases. The study integrates both structured data (e.g., demographics, lab results, medication history) and unstructured data (e.g., imaging reports, physician notes, ECG timestamps) to refine AI-guided dual antiplatelet therapy (DAPT) recommendations. While structured data offers measurable clinical variables, unstructured data boosts predictive accuracy through deep learning models. Regional variations in cardiovascular disease and treatment outcomes necessitate an adaptive, AI-driven framework to address both patient-specific and population-level differences. A cohort of 5,000 patients was analyzed, with 3,500 drawn from ICU-based registries (MIMIC-IV PhysioNet) and 1,500 from the UAE’s Bayanat Open Data Portal. This design ensures external validation across varied genetic backgrounds and treatment protocols, enhancing geographic and demographic applicability. The resulting risk prediction framework effectively balances ischemic and bleeding risks to personalize therapy. For a detailed report of all supplementary materials, see *Supplementary Files List*.

### 1. Medical Phenotypes and Endpoints

#### 1.1 Standardized Definitions and Reproducibility

Standardized endpoint definitions, widely accepted in cardiovascular research, were used to ensure clinical validity and reproducibility. Authenticated phenotypes improve the accuracy of AI-based risk stratification including Myocardial infarction, stroke or transient ischemic attack (TIA), definite/probable stent thrombosis, ischemic Endpoints (Major Adverse Cardiovascular Events, MACE) and cardiovascular mortality

- *Bleeding Endpoints (BARC Classification):*

- BARC Type 2: Bleeding requiring medical intervention

- BARC Types 3–5: Major complications, including fatal bleeding

These phenotypes were curated using ICD-10, SNOMED CT, and EHR clinical documentation to standardize classifications across datasets. AI-driven phenotypic stratification allows for dynamic DAPT adjustments tailored to individual risk profiles.

### 2. Risk Factors and Comorbidities

#### 2.1 Types of Risk Factors and Their Integration

The AI model incorporated diverse risk categories to enhance stratification by including: demographics ethnicity (Arab vs. non-Arab), age gender and geographic variations in prevalence, metabolic: obesity, diabetes and dyslipidemia, Cardiac Comorbidities: atrial fibrillation (AFib), prior ischemic heart disease (IHD), and congestive heart failure (CHF), and lifestyle modifications: Smoking (current, former) status. These variables were dynamically integrated to ensure ischemic protection was balanced without increasing bleeding risks.

### 3. Post-PCI Pharmacological Intervention

#### 3.1 DAPT Regimens and Adjunct Therapies

The study evaluated DAPT strategies, targeting ischemic and bleeding risk modulation for high-and low-risk groups, tailoring therapy duration based on AI-driven insights.

#### 3.2 Core Antithrombotic Combination Therapies

The Ticagrelor-ASA combination represents the antithrombotic regimen of choice for ACS patients without high bleeding risk, based on its established mortality benefit. Clopidogrel-Aspirin: Recommended alternative for bleeding-prone populations or carriers of loss-of-function CYP2C19 alleles (particularly *2 and *3 variants). Prasugrel-Aspirin: Optimal for STEMI patients <75 years without cerebrovascular disease (TRITON-TIMI 38: 19% RRR in primary endpoint, p<0.001).

#### 3.3 Concomitant Pharmacologic Modulators

The machine learning algorithm incorporated pharmacodynamic interactions from concomitant medications including: Cardiometabolic modulators (β-blockers, RAAS inhibitors), NSAIDs (competitive COX-1 inhibition), immunomodulators (corticosteroids), and direct oral anticoagulants (bleeding risk potentiation). This computational approach enabled real-time adjustment of thrombotic-hemorrhagic equilibrium predictions, facilitate: dynamic risk reclassification, patient-specific therapeutic intensity modulation, and temporal adjustments based on evolving pharmacologic profiles.

### 4. AI Model Development and Risk Stratification

#### 4.1 Machine Learning Models Applied for Event Prediction

Various machine learning techniques were utilized for training and validating risk prediction models, encompassing both primary and secondary analyses.

| Model | Primary Objective | Performance Metric |
| --- | --- | --- |
| Random Forest | Feature selection & ranking | SHAP Feature Importance |
| Neural Networks (ANNs) | Deep learning-based non-linear risk modeling | Calibration & Accuracy |
| LightGBM (Weighted LGBM) | Captures non-linear ischemic/bleeding risk interactions | AUROC & AUPRC |
| Logistic Regression | Baseline comparison | Concordance Index (C-Index) |

Through model comparisons, LightGBM emerged as the top performer, offering enhanced predictive accuracy and better interpretability compared to traditional approaches.

### 5. Data Preprocessing

The study integrated structured clinical data from multiple sources, including UAE electronic health records (demographics, comorbidities, lab values), PCI registries (stent type, SYNTAX scores), targeted genotyping (CYP2C19 polymorphisms), and lifestyle questionnaires (smoking, physical activity). Missing data were addressed through multiple imputation, while continuous variables were standardized (mean=0, SD=1) and outliers managed via IQR-based winsorization. Feature engineering created interaction terms (e.g., diabetes-stent type) and applied one-hot encoding for categorical variables to optimize machine learning compatibility.

### 6. AI Model Specifications

Three machine learning architectures were implemented: 1) Random Forest (500 trees, max depth=10), 2) Gradient Boosting Machines (learning rate=0.1, 1000 estimators), and 3) Deep Neural Networks (three-layer architecture with 128-64-32 neurons). Hyperparameter optimization employed grid search with 5-fold cross-validation. Model interpretability was ensured through SHAP values for global feature importance and LIME for local explanations, maintaining clinical transparency in predictions.

### 7. Model Validation & Clinical Integration

The dataset was split into training (70%) and testing (30%) sets, with performance assessed via AUC-ROC, F1-score, and precision-recall metrics. External validation used independent UAE clinical data, while expert review ensured clinical relevance. The finalized AI model was integrated into a CDSS, enabling dynamic DAPT adjustments based on real-time risk scores (ischemic threshold: 15%, bleeding threshold: 10%), calibrated via Youden’s Index. Genetic stratification revealed CYP2C19*2 carriers had 18-25% higher ischemic risk, necessitating ethnic-specific model adjustments for optimal personalization.

### 12. Supplementary Results

#### Supplementary Data and Transparency Statement

To ensure full transparency and facilitate clinical implementation, we provide comprehensive supplementary materials designed to meet FAIR (Findable, Accessible, Interoperable, Reusable) principles for AI in healthcare:

#### Data Tables

- Table S1: Final model variables with SHAP-derived clinical importance scores (stratified by ischemic/bleeding outcomes)
- Table S2: Cost-effectiveness model specifications (AED/QALY framework) with sensitivity analysis parameters

#### Visual Analytics

- Figure S1: Time-to-event analyses comparing UAE vs. global cohorts with BMI stratification
- Figure S2: Interactive SHAP waterfall plots demonstrating case-level prediction explainability.

#### Code Repository (Available via controlled access for validation studies)

- Data preprocessing pipeline (Python, scikit-learn) with EHR harmonization protocols
- Model training suites for all algorithms (LightGBM/RF/NN), including hyperparameter spaces
- Clinical deployment package:

Real-time SHAP explainer module
Calibration monitoring tools
EHR integration templates (HL7/FHIR compatible)

#### Justification for Supplementary Figures & Tables

To ensure consistency between the supplementary and primary analyses, the following measures were implemented:

- Tables S1–S8 were reviewed to maintain alignment in variable definitions and statistical reporting with the primary tables.

- Figures S1–S6 were cross-referenced with the primary figures to provide complementary insights without duplication.

- Supplementary Tables S4 and S6 were harmonized with primary Table 6 to ensure consistent reporting of AI performance metrics.

- Supplementary Table S5 now directly references primary Table 5, supporting the classification of ischemic and bleeding risks across various DAPT regimens.

#### Supplementary Tables

- **S1:** lists the final variables included in the AI prediction model, including SHAP score ranks for ischemic and bleeding risk stratification.
- **S2:** details the cost-effectiveness analysis model inputs and calculations in AED, aligning with QALY-based evaluations referenced in the manuscript.
- **S3:** lists the final variables included in the AI prediction model, including SHAP score ranks for ischemic and bleeding risk stratification.
- **S4:** Details the cost-effectiveness analysis model inputs and calculations in AED, aligning with QALY-based evaluations referenced in the manuscript.
- **S5:** Lists standardized medical codes for drug-eluting stent implantation, which is critical for linking procedural details to clinical outcomes.
- **S6:** Compares AI-DAPT models against baseline models, supporting the validity of AI-driven risk prediction.
- **S7:** Identifies the top 20 comorbidities influencing ischemic and bleeding risks, reinforcing the role of patient-specific risk stratification.
- **S8:** Presents model performance metrics, offering transparency on AI model efficacy beyond the primary analysis.
- **S9:** Evaluates survival differences between stratified groups, providing supplementary validation for Kaplan-Meier analysis.
- **S10:** Examines geographic and genetic polymorphism factors, highlighting regional variations in ischemic and bleeding risks.

#### Supplementary Figures

· **S1:** compares UAE-specific vs. global ischemic and bleeding risks across time intervals and BMI stratification.

- **S2:** presents SHAP-based feature importance rankings for both ischemic and bleeding outcomes.
- **S3:** Overlapping histograms for age and BMI distribution (Panel A), medication adherence rates (Panel B), and genetic risk score distributions (Panel C) validate key study variables.
- **S4:** Correlation heatmap highlights clinical and demographic associations with ischemic and bleeding outcomes.
- **S5:** AI model learning curves assess training performance, ensuring model stability and optimization.
- **S6:** Bar Chart Illustrating Ischemic and Bleeding Risk Prediction by Different DAPT Regimens Across Time Intervals (0-37 Months)

**Supplementary Table 1**: lists the final variables included in the AI prediction model, including SHAP score ranks for ischemic and bleeding risk stratification.

Interpretation & Significance

This table provides a comprehensive list of the clinical, demographic, procedural, and biochemical variables incorporated into the AI-driven prediction model. The selection of these variables was guided by:

- Domain expertise and clinical relevance,
- Evidence-based literature supporting their predictive value,
- Data availability across both the MIMIC-IV and UAE-EHR datasets.

Key features with high SHAP (SHapley Additive exPlanations) scores—such as DAPT duration, prior myocardial infarction, atrial fibrillation, chronic kidney disease, hemoglobin levels, and anticoagulant use—highlight the model’s reliance on clinically meaningful predictors.

**Supplementary Table 2:** details the cost-effectiveness analysis model inputs and calculations in AED, aligning with QALY-based evaluations referenced in the manuscript.

This analysis demonstrates AI-personalized DAPT’s economic dominance over fixed-duration therapy, with an ICER of -451,315 AED/QALY in the UAE healthcare context. Key drivers include: (1) optimized treatment durations reducing medication costs, and (2) fewer adverse events lowering hospitalization expenses. The model’s incorporation of regional cost structures and QALY adjustments enhances its real-world applicability. These findings provide robust evidence for implementing AI-driven DAPT personalization as both clinically and economically superior, offering policymakers a validated approach to enhance cardiovascular care efficiency while improving outcomes.

**Supplementary Table 3: Specific Codes and Types Used in Ischemic and Bleeding Endpoints**

Supplementary Table 3 details the standardized coding systems (ICD-10-CM, RxNorm, CPT, LOINC) used to define ischemic and bleeding endpoints. Ischemic events were captured through MI codes (I21.x, I22.x), cerebrovascular events (I63.x), PCI procedures (CPT 92928), and cardiac biomarkers (LOINC 32768-7). Bleeding events were identified via GI hemorrhage codes (K92.2), anemia (D62), and anticoagulant use (RxNorm 197361). Comorbidities (I10, E11.x) and DAPT medications (RxNorm 198211) were included for risk adjustment. This structured coding approach minimized misclassification while enabling precise pharmacotherapy and subgroup analyses.

**Supplementary Table 4: Dual Antiplatelet Therapy (DAPT) and Co-Medications used in Study**

The study evaluated key drug classes for post-PCI management:

1. DAPT therapeutics: P2Y12 inhibitors (clopidogrel/ticagrelor/prasugrel), combined with aspirin (COX-1 inhibitor) and ticagrelor favored for acute coronary syndrome owing for rapid onset.
2. Adjunctive treatments:

a. Anticoagulants (warfarin, DOACs like rivaroxaban) for patients requiring triple therapy
b. PPIs (pantoprazole favored with clopidogrel) for GI protection
c. Statins (atorvastatin/rosuvastatin) for atherosclerosis management
3. Comorbidity treatments:

SGLT2 inhibitors (empagliflozin) in diabetic cases
Beta-blockers (metoprolol/bisoprolol) in post-MI patients
ACE inhibitors (ramipril/lisinopril) for endothelial protection

This comprehensive pharmacological approach optimizes outcomes while minimizing risks in post-PCI care.

**Supplementary Table 5: Multisystem Coding Schema for Coronary Drug-Eluting Stents**

The following schema delineates DES categorization within standardized medical coding frameworks and contemporary PCI methodologies. Harmonized coding ontologies, including ICD-10-PCS, CPT, HCPCS, and RxNorm, provide granular procedural documentation and enhance clinical workflow optimization. **ICD-10-PCS procedural mapping:** Specific alphanumeric designations (027034Z, 02703DZ, 02713DZ) delineate DES deployment within percutaneous coronary revascularization. Current Procedural Terminology (CPT) descriptors (92928, 92933, 92937) classify coronary stent deployment, whereas HCPCS Level II identifiers (C9600-C9602) specify advanced polymer-based DES platforms, enabling precise procedural accounting and fiscal reconciliation.

### Generational Advancements in DES

Contemporary PCI utilizes generation-specific stent technologies: Second-generation EES/ZES platforms (RxNorm 691405/721306; LOINC 75934-1/82905-2) demonstrate improved safety profiles through chromium alloy architectures and biodegradable polymers. Third-generation innovations include fully bioresorbable scaffolds (ICD X2R034Z) and polymer-free designs (HCPCS C9607-8) that eliminate permanent implants. Specialized platforms address high-risk anatomies (02703ZZ) and comorbidities (027034Z/X2R034Y) through targeted drug-elution kinetics. Procedural optimization employs OCT (CPT 92978) for micron-level strut assessment and FFR (G0290) for ischemia quantification, enabling precision stent deployment.

**Supplementary Table 6:** Algorithmic Performance Comparison for Ischemic and Hemorrhagic Event Prediction in DAPT-Treated Cohorts

Benchmark analysis revealed conventional risk prediction models (logistic regression/Cox proportional hazards) employing standard clinical variables and DAPT scores provided limited discriminative accuracy (ischemia AUROC 0.68-0.72; bleeding 0.65-0.68) and suboptimal clinical precision (AUPRC 0.41-0.48), constraining their utility for personalized therapy. In contrast, our optimized XGBoost ensemble - integrating angiographic SYNTAX scores, high-sensitivity troponin trajectories, and longitudinal treatment adherence data - achieved significantly enhanced prediction (ischemia AUROC 0.86, 95% CI 0.83-0.89; bleeding 0.81, 0.78-0.84). The deep neural network architecture, processing multimodal inputs from critical care flowsheets and medication possession ratios, established new performance benchmarks (ischemia AUROC 0.89; bleeding 0.85), representing a 23-31% relative improvement over conventional methods. The optimized LightGBM framework with automated feature selection and temporal weighting emerged as the superior risk stratification tool, demonstrating both exceptional discrimination (AUROC 0.91/0.88) and clinical-actionable precision (AUPRC 0.83/0.79), representing a 32-41% improvement over conventional approaches.

**Supplementary Table 7:** Systematic Comorbidity Analysis for Precision Risk Assessment in Dual Antiplatelet Therapy Decision-Making

The comorbidity risk stratification analysis identified 20 clinically significant predictors of thrombotic and hemorrhagic events in DAPT-treated patients. Coronary artery disease emerged as the predominant ischemic risk determinant (risk coefficient 0.92, 95% CI 0.89-0.95), establishing atherosclerotic burden as the principal driver of thrombotic events. Atrial fibrillation demonstrated significant pleiotropic risk (ischemic coefficient 0.89; hemorrhagic 0.58), reflecting its dual pathophysiology of stasis-related thrombosis and anticoagulant sensitivity.

Diabetes mellitus exhibited substantial bidirectional risk potential (ischemic 0.87; hemorrhagic 0.52), highlighting the complex interplay between hypercoagulability and vascular fragility in metabolic syndrome. Renal impairment (coefficient 0.89) and prior hemorrhage (0.92) constituted the most potent hemorrhagic predictors, while hematologic (anemia 0.91) and hepatic dysfunction (0.89) represented multifactorial bleeding predispositions. Malignancy (bleeding coefficient 0.85) and heart failure (ischemic 0.78) exemplified the therapeutic dichotomy, with inflammatory-metabolic disorders (adiposity-related ischemic risk 0.71) further complicating risk stratification.

**Supplementary Table 8: Model Performance Metrics for AI-Based Risk Prediction**

Weighted LightGBM demonstrated superior predictive capability, achieving the highest AUROCs (0.88 ischemic, 0.85 bleeding) and AUPRCs (0.74 ischemic, 0.71 bleeding), along with optimal calibration (0.91) and F1-scores (0.82). Neural networks and random forests showed competitive ischemic risk prediction (AUROCs 0.86-0.87) but were less robust for bleeding events. Traditional methods (logistic regression: AUROC 0.78-0.80; SVM) underperformed across all metrics, particularly in precision-recall balance (AUPRCs 0.60-0.65). These results establish LightGBM as the optimal model for DAPT risk stratification, combining advanced feature selection with temporal modeling to enhance clinical decision-making.

**Supplementary Table 9: Log-Rank Test Results Comparing Survival Between Stratified Groups**

AI-personalized DAPT demonstrated superior outcomes with the highest median survival (45.8 months, 95% CI 43.6–48.1) and event-free survival rate (91.5%), serving as the reference group. While extended DAPT (≥24 months) showed comparable results (40.2 months, p=0.073; HR 1.10), standard 12-month DAPT had significantly worse outcomes (34.7 months, p=0.017; HR 1.23). Short-duration DAPT (<6 months) performed poorest (24.5 months, p=0.0012; HR 1.58) with only 72.3% event-free survival. These findings confirm AI-guided personalization optimizes survival benefits while minimizing adverse events compared to fixed-duration approaches.

**Supplementary Table 10:** Multidimensional Risk Profiling: Geographic, Pharmacogenetic, and Lifestyle Predictors of Thrombotic and Bleeding Outcomes

1. **Geospatial**: Urban residency predicted 35% greater ischemic risk (aHR 1.35, *p*=0.008), with 4.5% absolute risk disparity.
2. **Genetic**: *CYP2C19*2 alleles conferred 67% higher ischemic risk (aHR 1.67, *p*=0.001; PAR 21.5%), while *CYP2C19*17 variants increased bleeding risk by 39% (*p*=0.012).
3. **Ethnic**: Arab populations exhibited 54% elevated ischemic risk (aHR 1.54, *p*=0.006) with 17.8% population-attributable risk.
4. **Environmental**: Air pollution (aHR 1.41, *p*=0.011; PAR 13.9%) and sedentarism (ischemic aHR 1.32) were significant modifiable factors.

**Supplementary Figure 1:** The analysis reveals key temporal risk patterns: ischemic events peak at 6-12 months (post-DAPT cessation) while bleeding risks escalate at 12-18 months (with prolonged therapy). UAE-specific data highlight obesity’s disproportionate impact on ischemic risk compared to global cohorts.

In the AI-stratified cohort (n=3,100/5,000):

- 1,100 high bleeding-risk patients received shortened DAPT
- 2,400 high ischemic-risk patients continued extended therapy
- 1,800 achieved optimized durations through AI guidance

These results demonstrate AI’s capacity to dynamically balance ischemic/bleeding risks by personalizing DAPT duration.

**Supplementary Figure 2:** The analysis revealed distinct predictive hierarchies:

I**schemic risk**: prior MI (SHAP value 0.92), diabetes (0.87), and CKD (0.85) were dominant, alongside modifiable factors like DAPT duration (0.89) and stent type (0.82).

**Bleeding risk:** was primarily driven by age (0.91), anemia (0.89), anticoagulant use (0.85), and prior bleeding (0.83).

Key overlapping factors (atrial fibrillation, CKD) highlighted the challenge of dual-risk balance. The model’s identification of both established and novel (UAE-specific) predictors enhances its clinical utility for personalized DAPT decisions.

**Supplementary Figure 3: (a) Overlapping Histograms of Age and BMI Distributions in the Patient Cohort. (b) Comparative Distribution of Medication Adherence. (C) Genetic Risk Scores in the Study Cohort**

Panel (a): Population Characteristics Visualization: Bivariate analysis reveals: (i) Right-skewed age distribution (median 61 years, IQR 54-67) with peak density at 55-65 years (n=220); (ii) BMI distribution showing significant positive skew (γ =1.8) with obesity-class I predominance (28-32 kg/m², n=180). The age-BMI covariance structure (Mahalanobis D²=4.7, p<0.001) confirms obesity as a time-dependent risk modulator, consistent with our random forest feature ranking (mean decrease accuracy=2.3%) and temporal trajectory analysis (Supplementary Figure 3). Panel (b): Therapeutic Adherence Patterns: Bimodal pharmacotherapy compliance emerges with: (i) Primary peak at suboptimal 60-70% adherence (n=210, 36.8%); (ii) Secondary peak at therapeutic target (≥90%, n=160). The low-adherence cohort (<50%, n=80) demonstrates significantly reduced event-free survival (log-rank χ²=24.7, p<0.001), validating AI-derived adherence thresholds (Youden’s index=0.61) for ischemic risk prediction (AUC=0.79, 95% CI 0.73-0.85). Panel (c): The genetic risk score (GRS) is concentrated at 0.4–0.6 (250 patients, 50%), indicating moderate ischemic risk. A smaller subset (GRS > 0.7, 80 patients, 16%) faces elevated cardiovascular risk, requiring extended DAPT. Combining these findings with Panel B reveals that low adherence among high-GRS patients amplifies ischemic vulnerability, reinforcing AI’s importance in precision therapy.

**Supplementary Figure 4: Correlation Heat map of Clinical and Demographic Parameters for Post-PCI Risk Prediction**

This heatmap demonstrates correlations among key clinical and demographic factors using a gradient scale, with strong positive correlations in red and negative ones in blue. Quantitative correlation modeling demonstrated moderate covariance between chronological age and BMI (β=0.45, p<0.01), establishing adiposity accumulation as a time-dependent metabolic risk variable. Strong linear dependence was observed between elevated resting heart rate and ischemic complications (Spearman’s ρ=0.72, p<0.001), validating RHR as a continuous risk stratifier in PCI populations. Survival analysis demonstrated strong negative association between therapeutic adherence and event-free survival (Kendall’s τ=-0.61, p<0.001), establishing pharmacologic compliance as a modifiable protective factor.The artificial intelligence feature selection demonstrates criterion validity through significant genomic-phenotypic correlation (GRS-ischemia r=0.61, p<0.01), with corresponding Kaplan-Meier (Figure 2), estimates showing clear risk stratification (30-day event-free survival 82.4% vs 63.1%, p<0.001). Weak correlation between BMI and bleeding events (r = 0.28, p = 0.03) suggests obesity impacts ischemic risks more than bleeding, underscoring the need for dynamic risk stratification (Figure 6A). These findings refine AI-based decision-making to enhance post-PCI outcomes and precision treatment strategies.

**Supplementary Figure 5: Learning Curves for AI Model Development**

This figure displays AI model learning curves, showing the relationship between epochs and loss reduction during training. The training loss (blue) starts at 0.72, decreasing steadily to 0.12 by 40 epochs, indicating successful convergence. Validation loss (orange) follows a similar trend, stabilizing at 0.18 but diverging slightly after 35 epochs, suggesting potential overfitting or data variability.

- A sharp loss decline in the first 10 epochs reflects the model’s rapid initial learning phase.

- A minimal gap (0.06) between training and validation losses at final epochs demonstrates strong generalization and robustness for ischemic and bleeding risk predictions.

- Significant validation loss increases would have indicated overfitting, requiring adjustments like dropout layers or early stopping.

The figure confirms the AI model’s ability to capture patient-specific risk factors for DAPT personalization. The steady convergence of loss metrics ensures predictive stability, reinforcing its reliability in dynamic risk assessment (Figure 6). These trends align with high AUC values in ROC analysis (Figure 1B), validating the model’s readiness for clinical post-PCI applications.

**Supplementary Figure 6: Bar Chart Illustrating Ischemic and Bleeding Risk Prediction by Different DAPT Regimens Across Time Intervals (0-37 Months.**

This bar chart compares ischemic and bleeding risk predictions for Ticagrelor-only and Ticagrelor + Aspirin regimens over time.

### Ischemic Risk Prediction

- At 6 months, ischemic risk is low (8% for Ticagrelor-only, 5% for dual therapy), reflecting effective platelet inhibition.

- By 12 months, the risk increases to 12% in Ticagrelor-only but remains 8% with dual therapy, showing the added benefit of combination treatment.

- At 24 months, ischemic risk rises to 20% in Ticagrelor-only and 14% in dual therapy, emphasizing extended ischemic protection with dual therapy.

### Bleeding Risk Prediction

- At 6 months, prolonged DAPT exhibited time-dependent bleeding hazard (24-month ARD 6.3%, p=0.003)

- By 12 months, cumulative bleeding incidence diverged further (10.2% [8.7-11.9] dual therapy vs 6.1% [4.9-7.5] monotherapy, p<0.001), with number needed to harm (NNH) of 25 for continued dual therapy.

- At 24 months, bleeding events reach 18% with dual therapy compared to 12% in Ticagrelor-only, indicating heightened susceptibility from prolonged dual therapy.

### Pharmacodynamic and Outcomes Analysis Reveals

- While ticagrelor monotherapy demonstrates superior safety, its attenuated antiplatelet effect may compromise ischemic protection in high-risk patients (24-month MI rates: 3.8% vs 2.1%, p=0.02).

- Dual therapy maintains superior ischemic protection (24-month MI reduction 38%, p=0.007) but exacts a bleeding toll that escalates with treatment duration (12-month BARC ≥2 bleeding: 10.2% vs 6.1%).

### Study Integration and Conclusion

Aligned with Kaplan-Meier (Figure 1A) and Time-to-Event analyses (Figure 4A), these results validate AI-driven personalization of DAPT regimens. Precision adjustments ensure optimal protection, balancing ischemic and bleeding risks to improve post-PCI outcomes.

## 13. Limitations

### 13.1 Data Quality Challenges

Despite preprocessing, the study faced issues with missing values and data inconsistencies. Although multiple imputation techniques addressed gaps, imputed values might not fully reflect true variability. Using retrospective electronic health records (EHRs) included additional challenges like partial documentation and inconsistent data gathering across centers. These limitations highpoint the essential for prospective studies with homogenous protocols to advance data consistency.

### 13.2 Limitations in External Validity

While the analytical model established robust performance in the UAE cohort, its generalizability to deviating populations needs cautious consideration. Region-specific features - including the elevated incidence of metabolic conditions (particularly diabetes mellitus at 38.7%) and population-specific genetic variants (e.g., Arab-specific polymorphisms in CYP2C19) - might constrain direct extrapolation. Furthermore, institutional variations in treatment protocols and healthcare delivery systems necessitate multinational validation across diverse care environments to establish universal applicability.

### 13.3 Clinical Integration Barriers

Real-world clinical incorporation meets multiple trials: (1) practitioner worry restricting from algorithmic misunderstandings and model interpretability restrictions; (2) systemic infrastructure deficits; and (3) ethical-legal complications concerning health data governance. These factors mutually contribute to therapeutics in implementation patterns. Effective integration into workflow involves robust IT set-up and interoperability with EHR systems, which are not generally obtainable. Addressing these barriers calls for clinician training, infrastructure investment, and user-friendly AI interfaces to facilitate adoption. By tackling these challenges, the framework’s potential for dynamic, AI-driven DAPT management can be fully realized, ensuring higher-quality and patient-centered care across diverse settings.

## 14. Perspective: Emerging Research Priorities

These results provide a scaffold for addressing three key unmet needs in cardiovascular risk prediction.

### 14.1 Wearable Integration

Future investigative priorities should focus on three key domains: (1) technical validation of consumer wearable data against gold-standard measures, (2) development of robust algorithms for artifact detection and signal processing in free-living environments, and (3) prospective evaluation of clinical utility through randomized implementation studies. Particular emphasis should be placed on establishing whether continuous physiological monitoring improves upon traditional risk stratification approaches in both sensitivity and specificity for adverse cardiovascular events.

### 14.2 Federated Artificial Intelligence

Distributed collaborative learning overcomes external validity constraints through institutionally partitioned model development across heterogeneous cohorts while maintaining data confidentiality. This paradigm enhances model robustness across populations and addresses critical data protection requirements, demonstrating particular utility for multi-center clinical validation studies. The federated artificial intelligence approach resolves limitations in cross-population applicability by enabling confidential model training across distributed datasets, simultaneously enhancing clinical utility while complying with stringent data governance requirements.

### 14.3 Extended follow-up measures

While this investigation focused on acute outcomes, longitudinal assessment of survival analyses, major adverse cardiovascular event (MACE) recurrence, and patient-reported outcomes remains essential for comprehensive evaluation of algorithm-guided DAPT optimization.

Prospective cohort studies incorporating extended surveillance periods (≥5 years) should investigate the temporal stability of computational recommendations and their sustained effects on composite clinical endpoints and health status metrics. This refined research agenda identifies pivotal knowledge gaps in post-PCI management while advancing the paradigm of phenotype-specific antithrombotic therapy optimization.

## 15 How and why Future Researchers could address These Points?

### 1. Wearable Integration

Why: Wearables offer real-time data to enhance accuracy and adaptiveness in DAPT strategies, enabling personalized, dynamic risk management.

How: Collaborate with wearable technology companies to create algorithms that integrate wearable metrics into AI models. Conduct pilot studies to evaluate the feasibility and impact of wearables on clinical outcomes.

### 2. Federated Learning

Why: Federated learning overcomes data privacy and generalizability issues, offering a scalable approach for multi-center collaborations.

How: Build federated learning networks with healthcare centers, adhering to strict data privacy protocols. Develop standardized agreements for consistent data collection and seamless collaboration.

### 3. Longitudinal clinical endpoints

Extended clinical surveillance through composite endpoints (mortality, functional status, and healthcare utilization) provides multidimensional evaluation of algorithmic decision support. The study design is conducted using multicenter longitudinal observational studies featuring extended surveillance periods (≥3 years), composite primary endpoints, and mixed-methods outcome assessment. The research framework: Establish registry-based studies showing periodic outcome assessments, landmark analysis at 12, 24, and 36 months, and embedded PRO collection protocols. Operationalization: Develop strategic partnerships with learning healthcare systems for pragmatic trial implementation, disease-specific registries for population-based sampling and community-based organizations for diverse population inclusion. And the study conducted establish multidisciplinary consortia incorporating health system partners for real-world data infrastructure, patient stakeholder groups for outcome relevance, and research networks for methodological rigor.

