## Supplementary Table S1- S10 for "AI-Driven Personalization of Dual Antiplatelet Therapy Duration Post-PCI: A Novel Approach Balancing Ischemic and Bleeding Risks"

### Slide 1
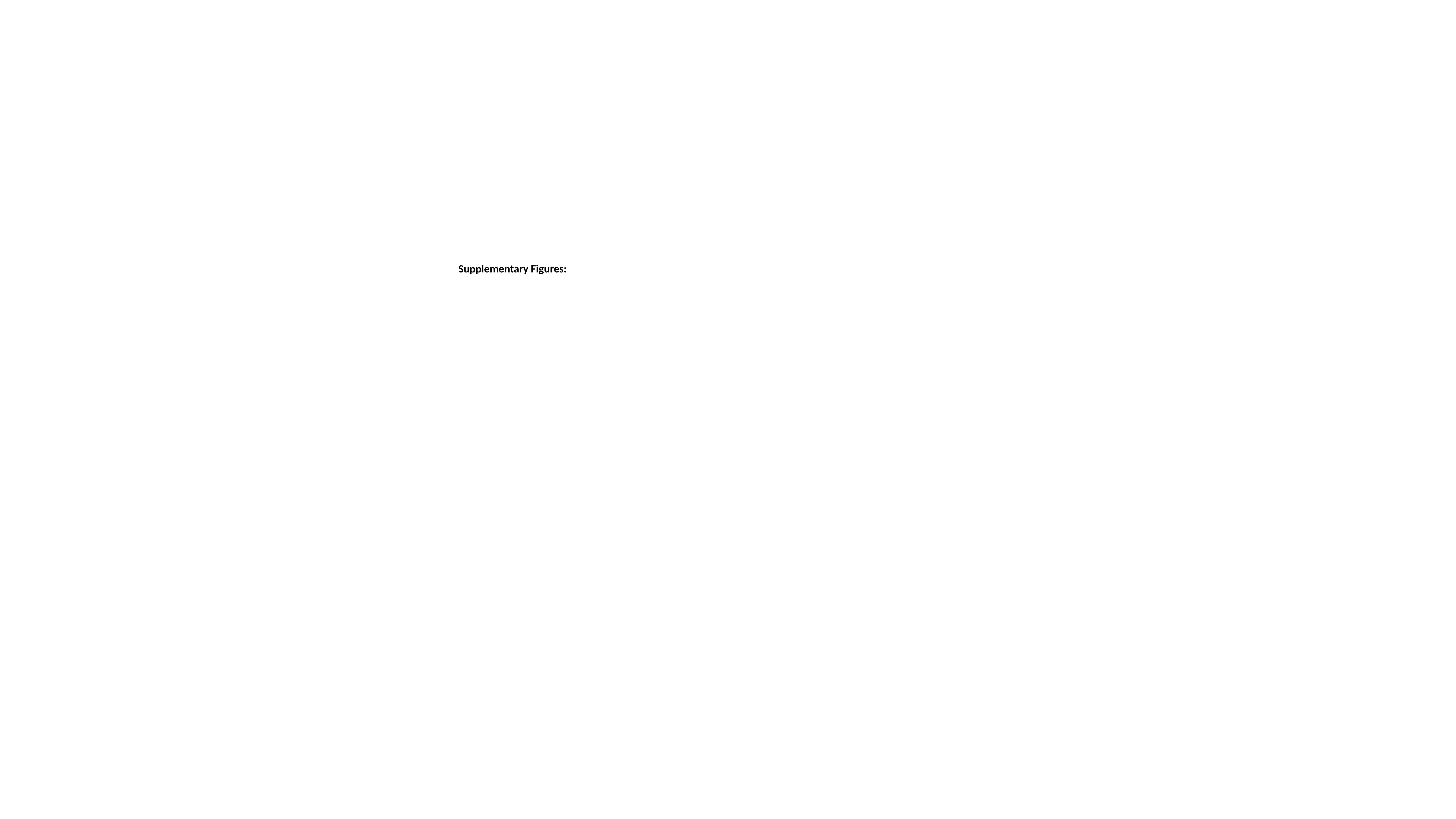

Supplementary Figures:

### Slide 2
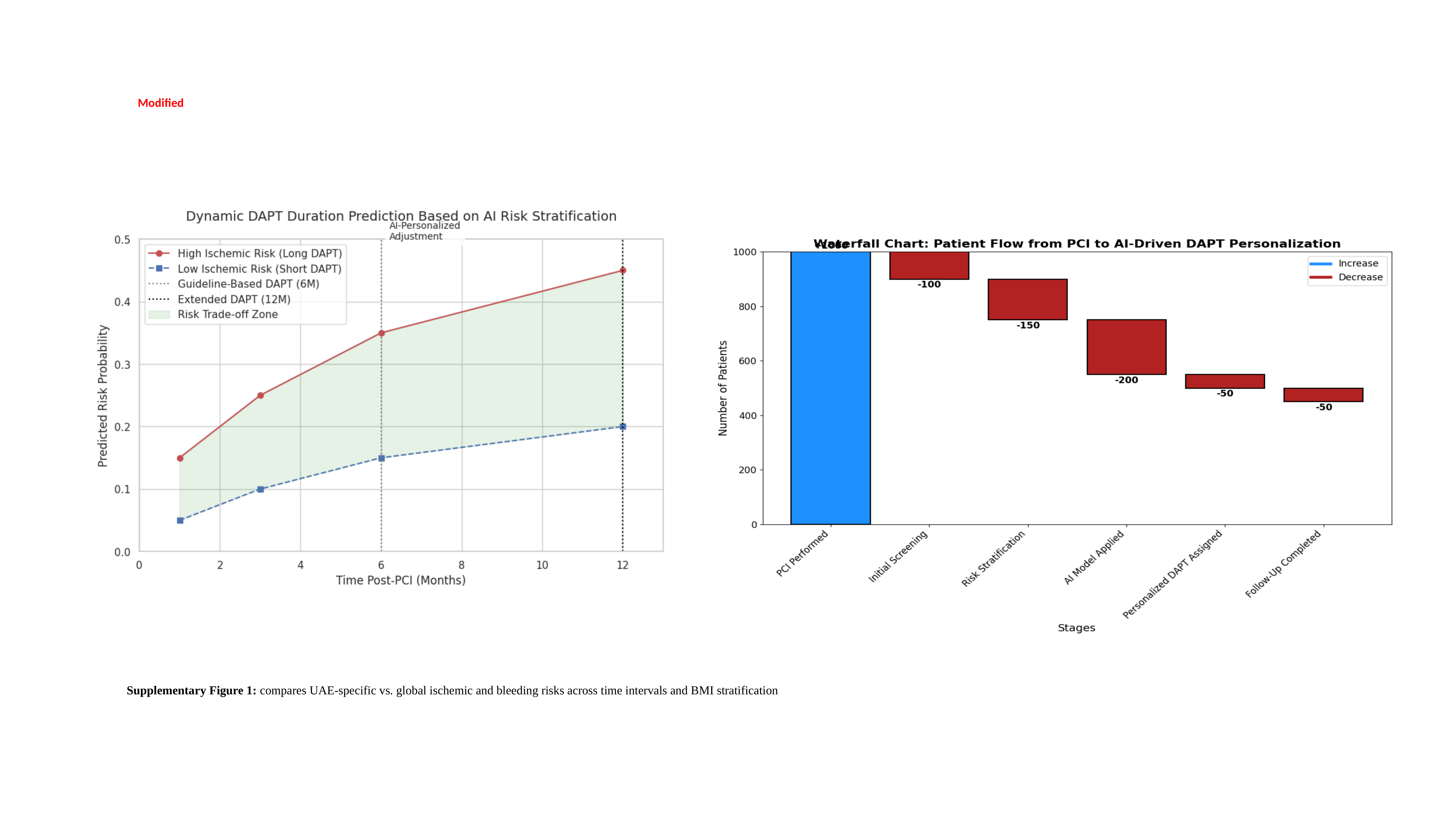

Modified
Supplementary Figure 1: compares UAE-specific vs. global ischemic and bleeding risks across time intervals and BMI stratification

### Slide 3
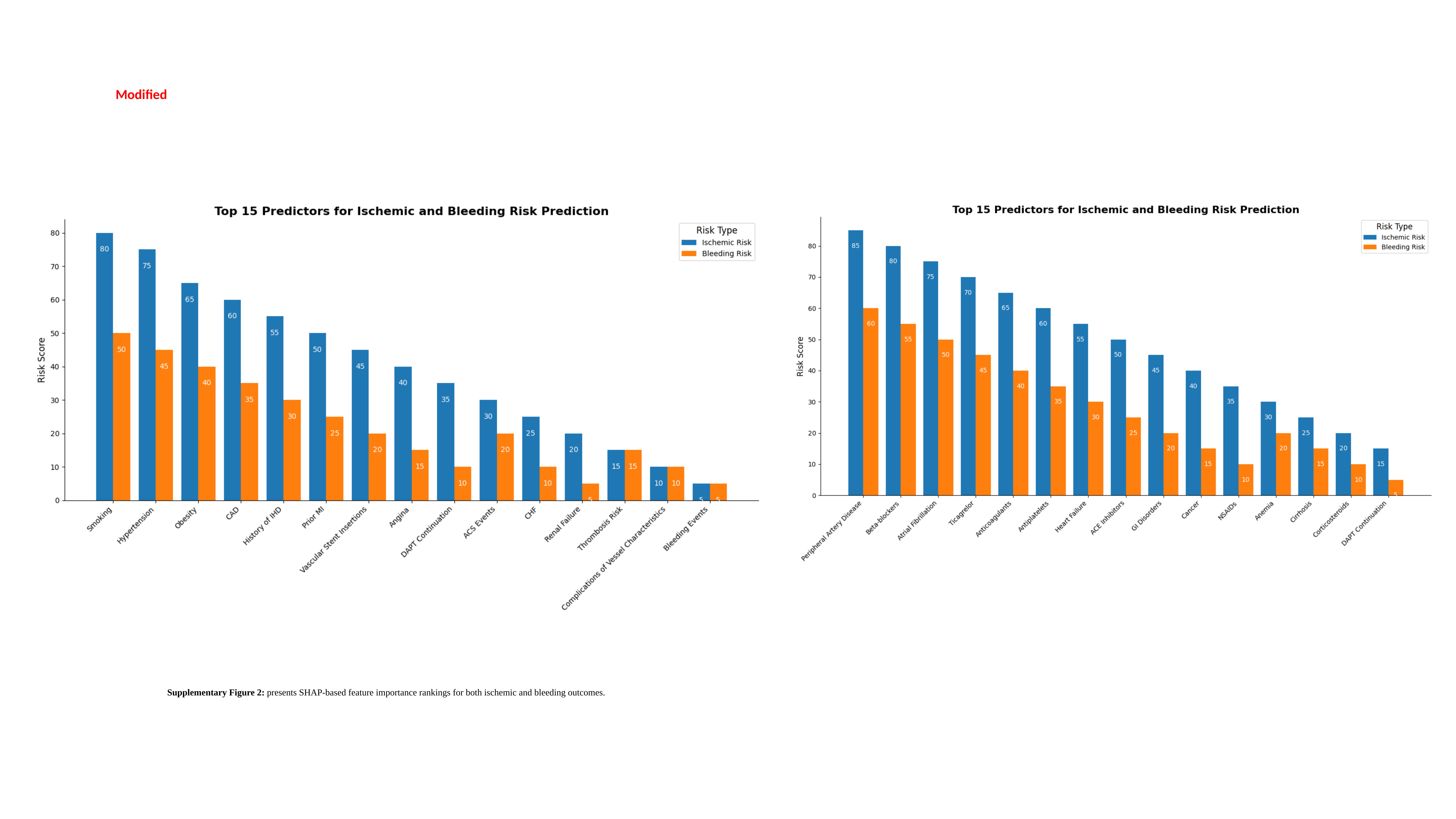

Modified
Supplementary Figure 2: presents SHAP-based feature importance rankings for both ischemic and bleeding outcomes.

### Slide 4
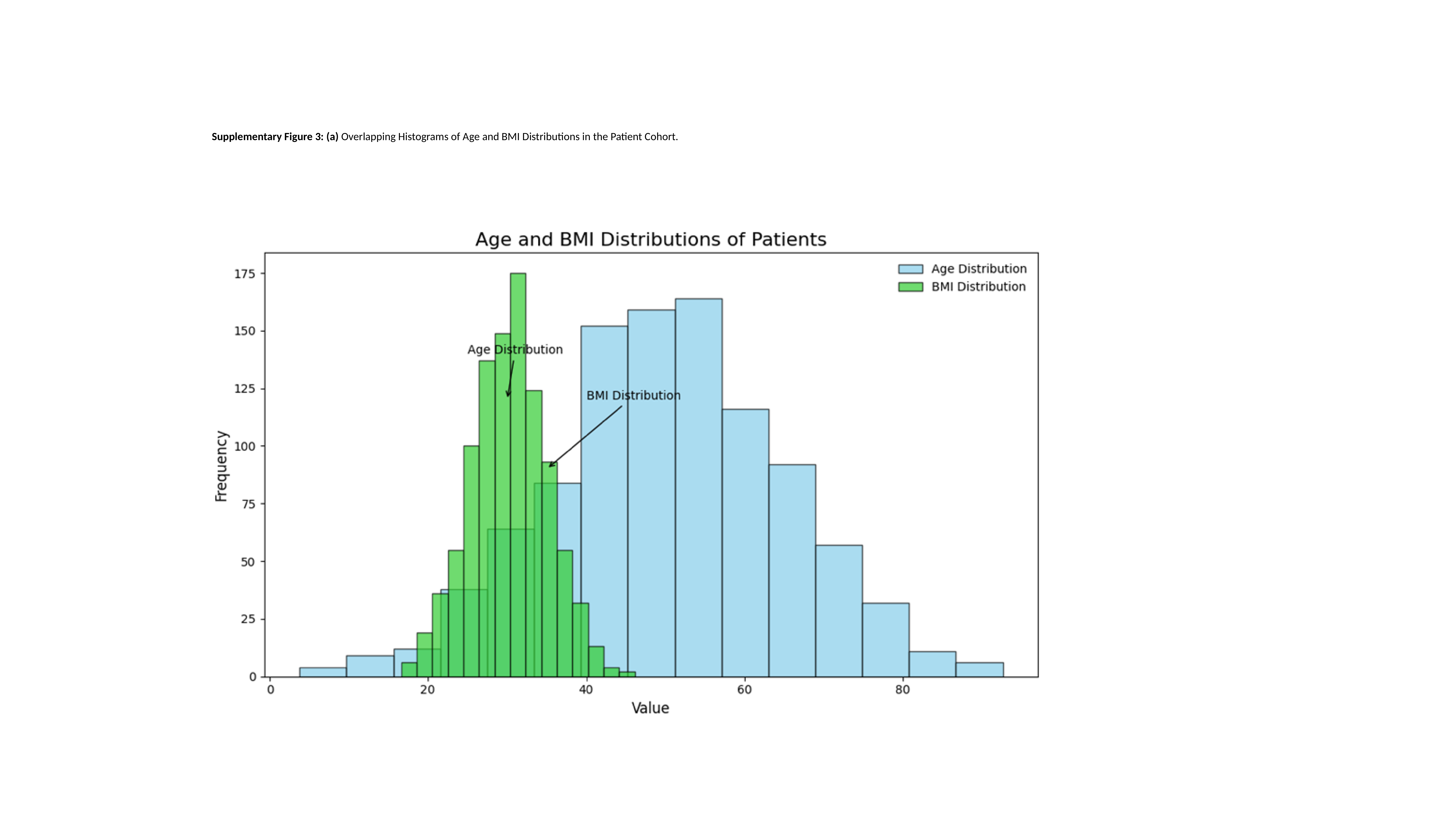

Supplementary Figure 3: (a) Overlapping Histograms of Age and BMI Distributions in the Patient Cohort.

### Slide 5
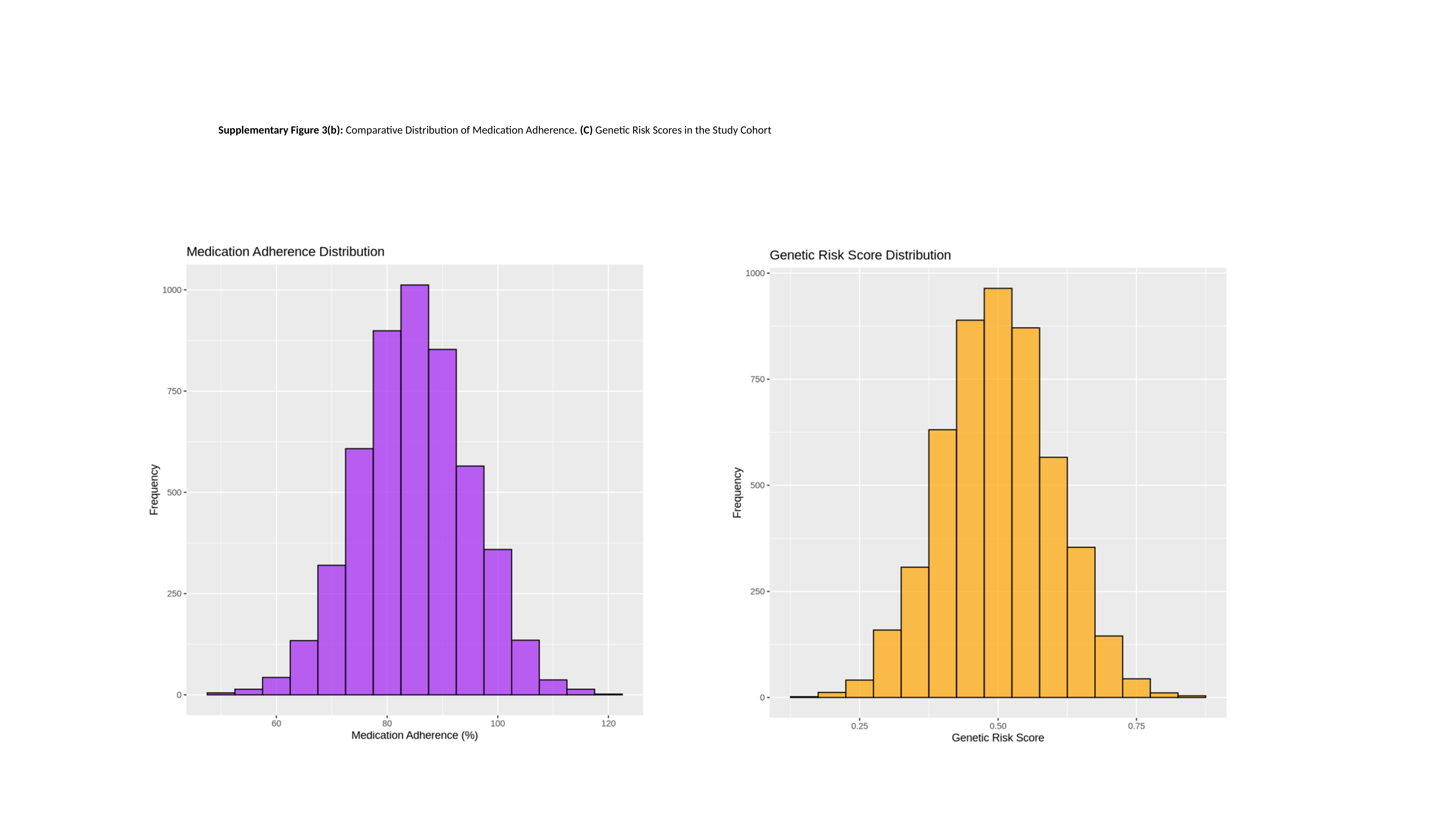

Supplementary Figure 3(b): Comparative Distribution of Medication Adherence. (C) Genetic Risk Scores in the Study Cohort

### Slide 6
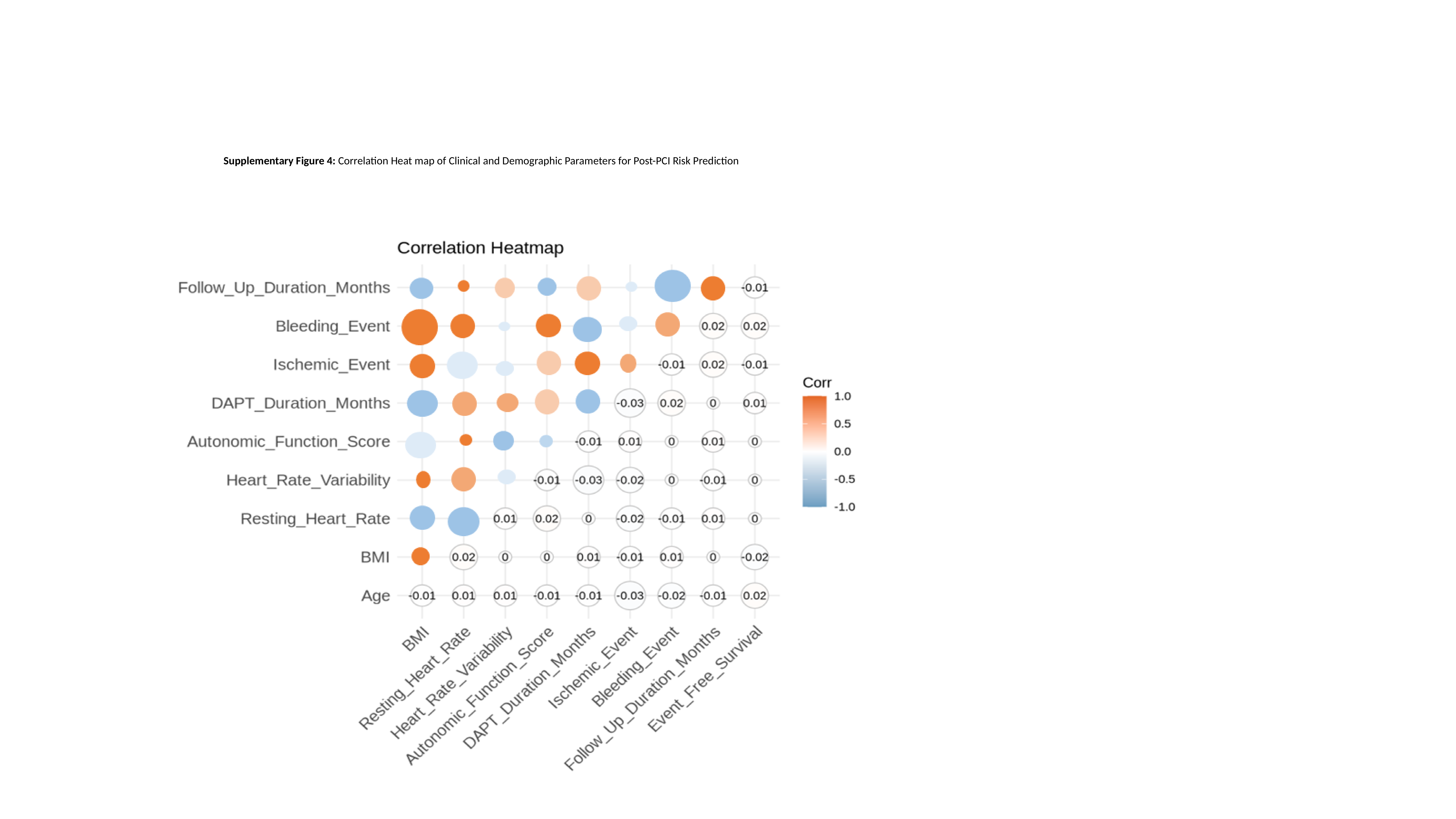

Supplementary Figure 4: Correlation Heat map of Clinical and Demographic Parameters for Post-PCI Risk Prediction

### Slide 7
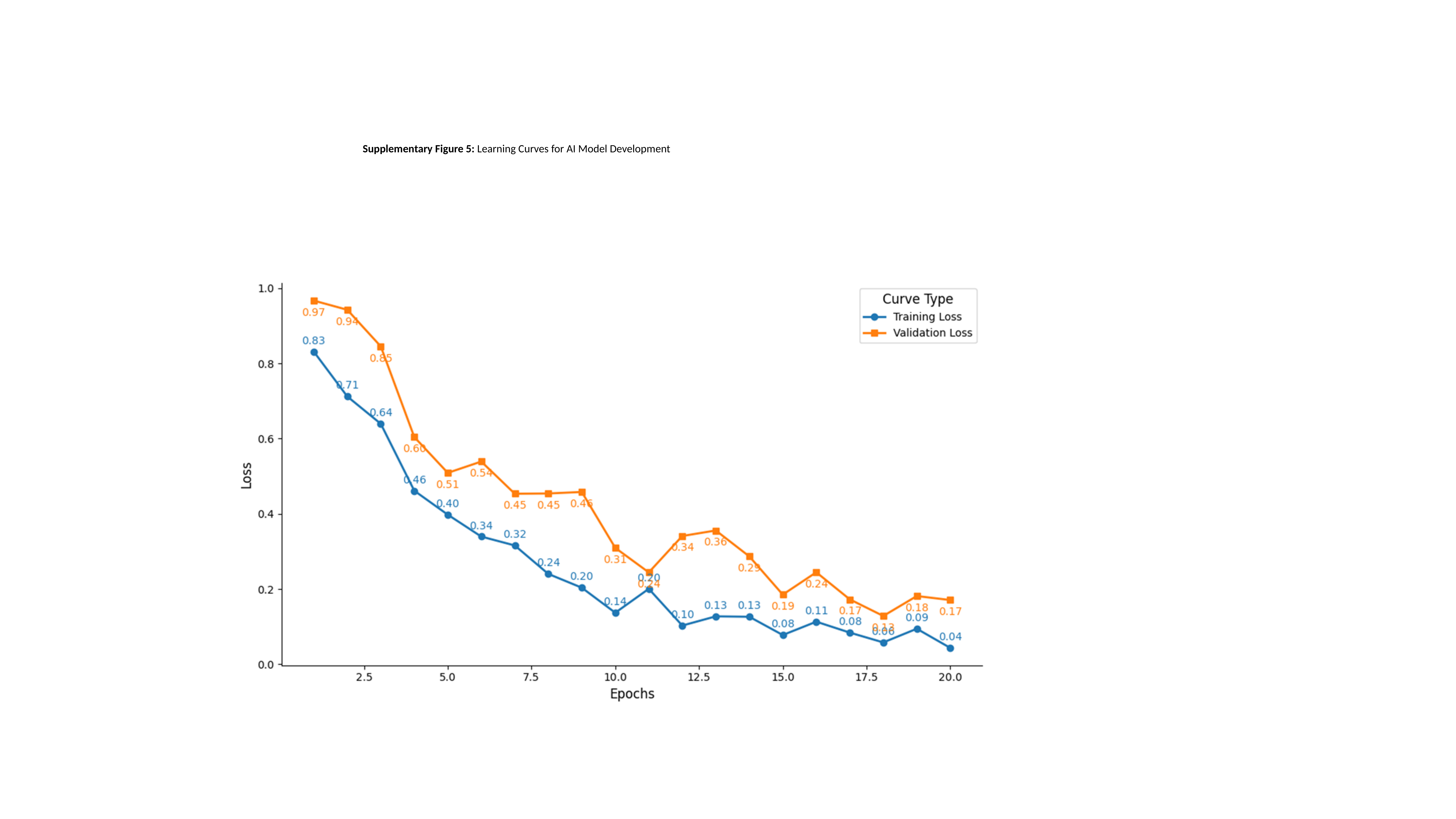

Supplementary Figure 5: Learning Curves for AI Model Development

### Slide 8
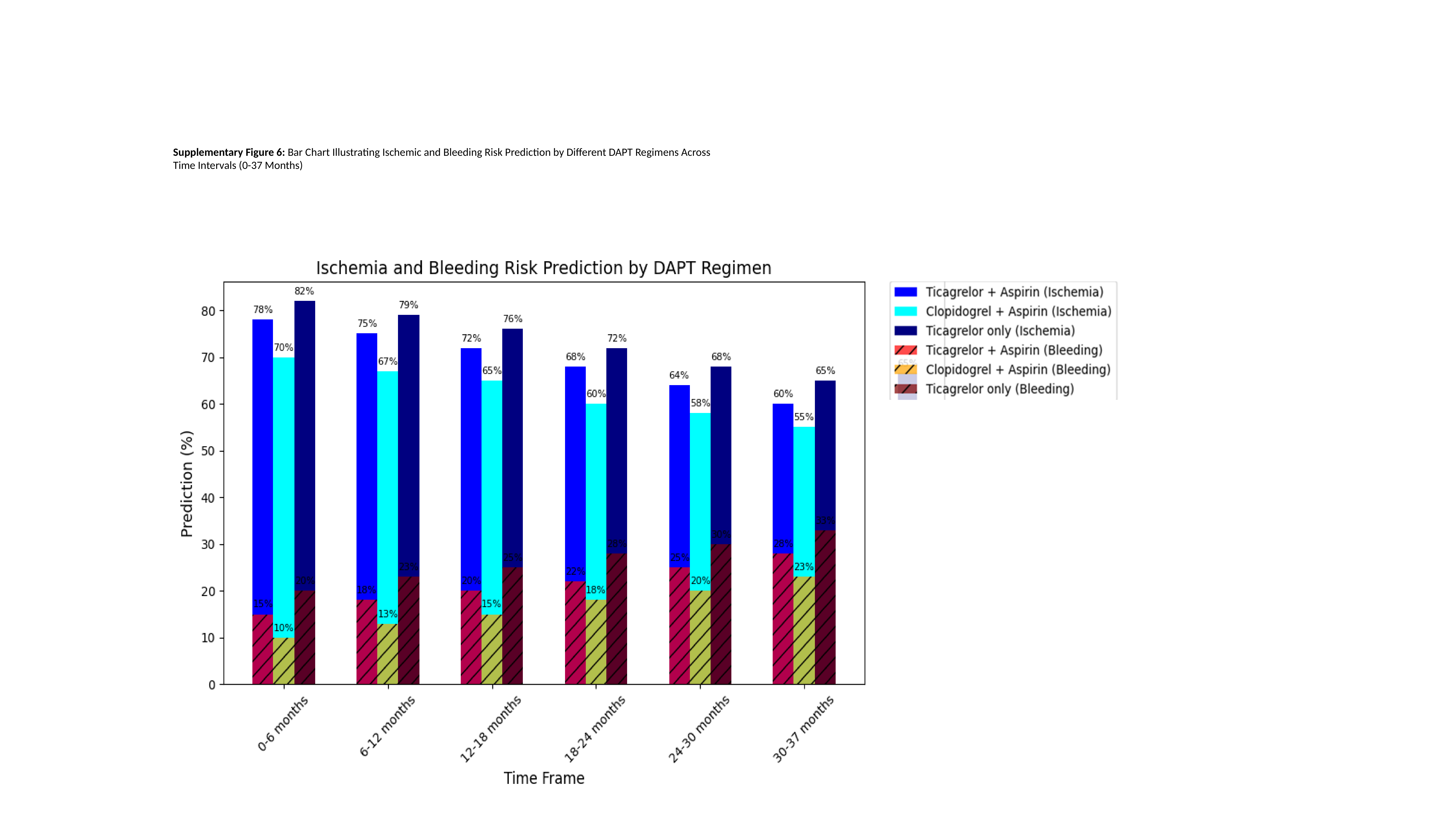

Supplementary Figure 6: Bar Chart Illustrating Ischemic and Bleeding Risk Prediction by Different DAPT Regimens Across Time Intervals (0-37 Months)

### Slide 9
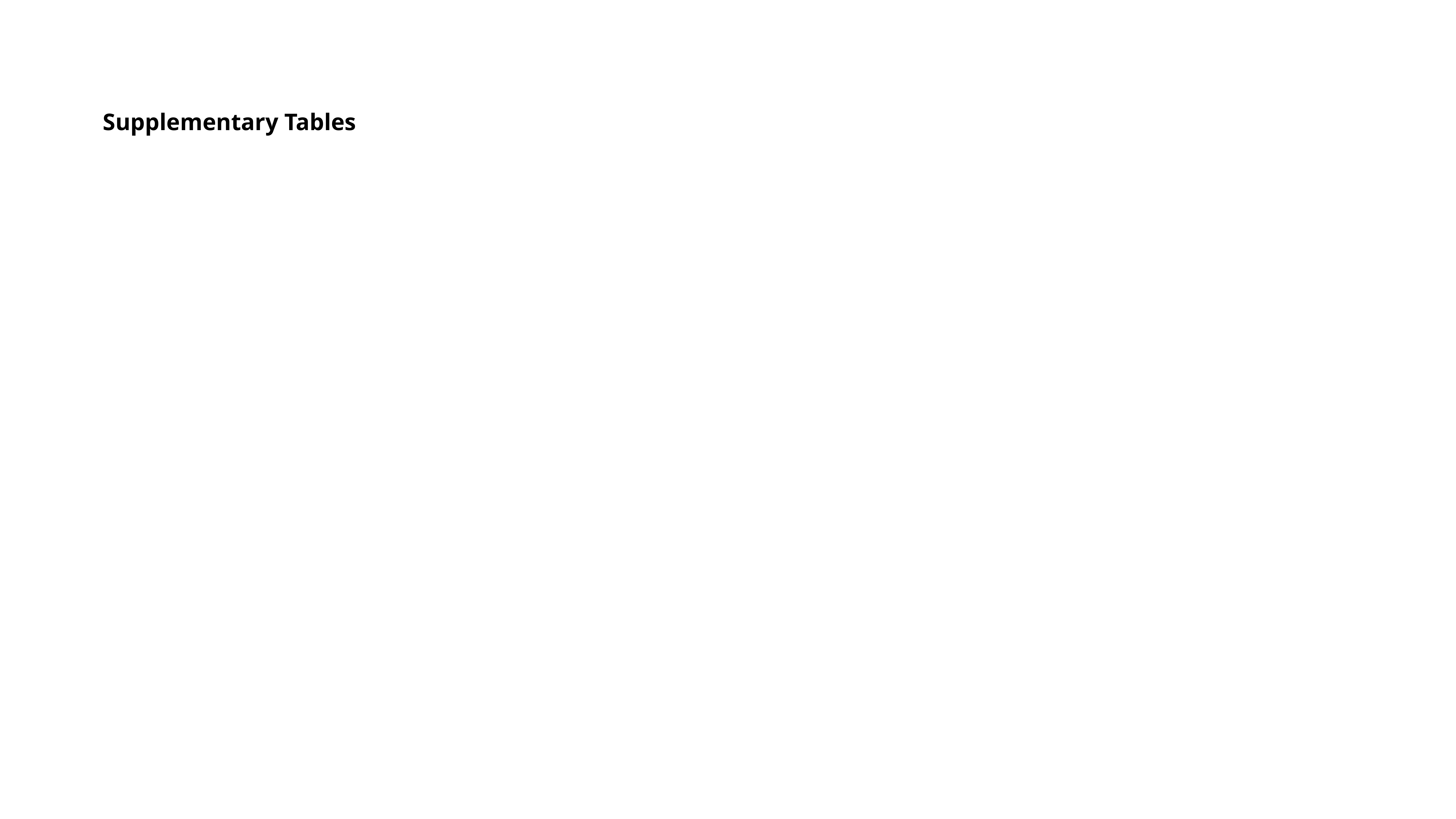

#### Supplementary Tables

### Slide 10
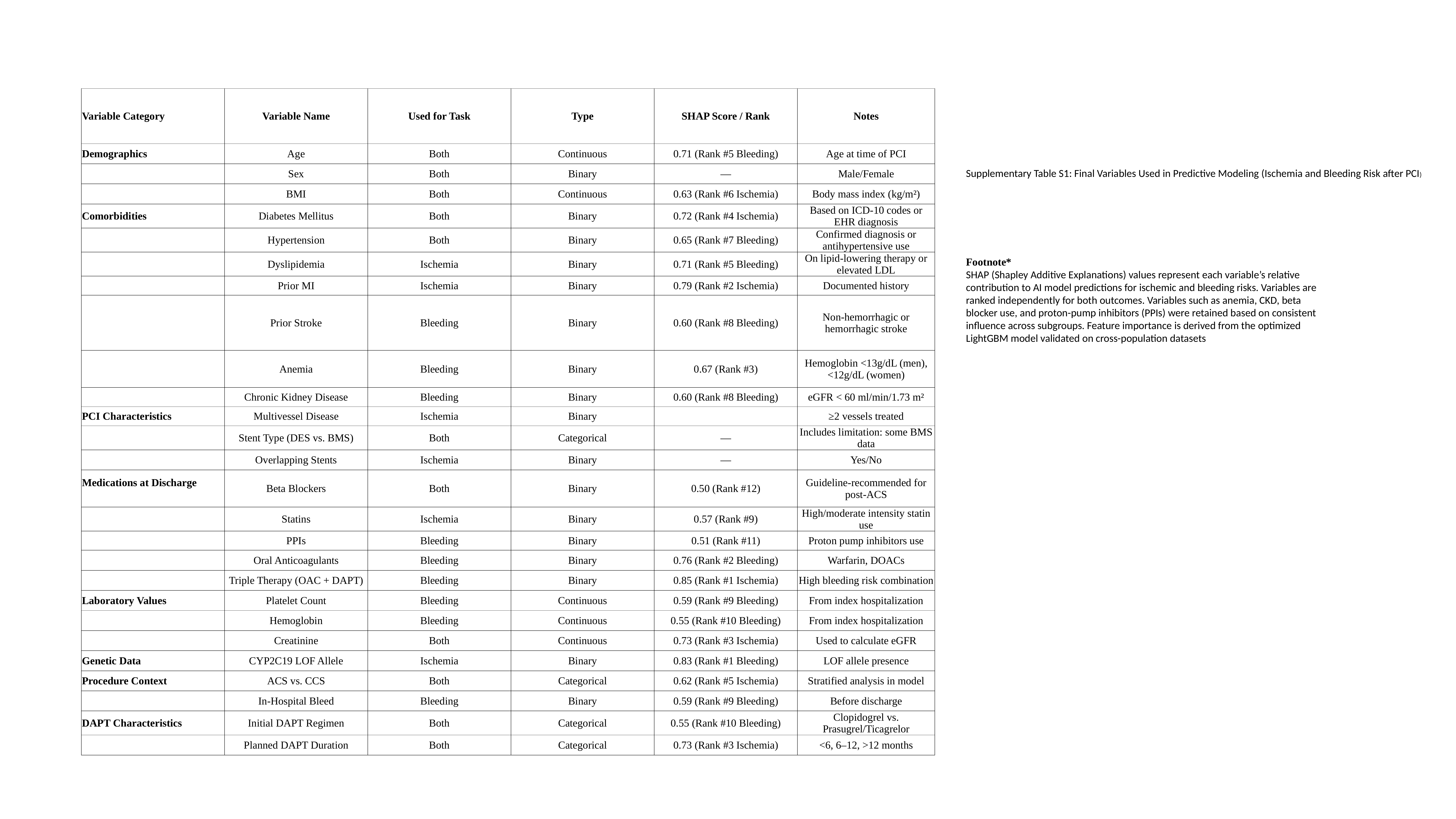

| Variable Category | Variable Name | Used for Task | Type | SHAP Score / Rank | Notes |
| --- | --- | --- | --- | --- | --- |
| Demographics | Age | Both | Continuous | 0.71 (Rank #5 Bleeding) | Age at time of PCI |
| | Sex | Both | Binary | — | Male/Female |
| | BMI | Both | Continuous | 0.63 (Rank #6 Ischemia) | Body mass index (kg/m²) |
| Comorbidities | Diabetes Mellitus | Both | Binary | 0.72 (Rank #4 Ischemia) | Based on ICD-10 codes or EHR diagnosis |
| | Hypertension | Both | Binary | 0.65 (Rank #7 Bleeding) | Confirmed diagnosis or antihypertensive use |
| | Dyslipidemia | Ischemia | Binary | 0.71 (Rank #5 Bleeding) | On lipid-lowering therapy or elevated LDL |
| | Prior MI | Ischemia | Binary | 0.79 (Rank #2 Ischemia) | Documented history |
| | Prior Stroke | Bleeding | Binary | 0.60 (Rank #8 Bleeding) | Non-hemorrhagic or hemorrhagic stroke |
| | Anemia | Bleeding | Binary | 0.67 (Rank #3) | Hemoglobin <13g/dL (men), <12g/dL (women) |
| | Chronic Kidney Disease | Bleeding | Binary | 0.60 (Rank #8 Bleeding) | eGFR < 60 ml/min/1.73 m² |
| PCI Characteristics | Multivessel Disease | Ischemia | Binary | | ≥2 vessels treated |
| | Stent Type (DES vs. BMS) | Both | Categorical | — | Includes limitation: some BMS data |
| | Overlapping Stents | Ischemia | Binary | — | Yes/No |
| Medications at Discharge | Beta Blockers | Both | Binary | 0.50 (Rank #12) | Guideline-recommended for post-ACS |
| | Statins | Ischemia | Binary | 0.57 (Rank #9) | High/moderate intensity statin use |
| | PPIs | Bleeding | Binary | 0.51 (Rank #11) | Proton pump inhibitors use |
| | Oral Anticoagulants | Bleeding | Binary | 0.76 (Rank #2 Bleeding) | Warfarin, DOACs |
| | Triple Therapy (OAC + DAPT) | Bleeding | Binary | 0.85 (Rank #1 Ischemia) | High bleeding risk combination |
| Laboratory Values | Platelet Count | Bleeding | Continuous | 0.59 (Rank #9 Bleeding) | From index hospitalization |
| | Hemoglobin | Bleeding | Continuous | 0.55 (Rank #10 Bleeding) | From index hospitalization |
| | Creatinine | Both | Continuous | 0.73 (Rank #3 Ischemia) | Used to calculate eGFR |
| Genetic Data | CYP2C19 LOF Allele | Ischemia | Binary | 0.83 (Rank #1 Bleeding) | LOF allele presence |
| Procedure Context | ACS vs. CCS | Both | Categorical | 0.62 (Rank #5 Ischemia) | Stratified analysis in model |
| | In-Hospital Bleed | Bleeding | Binary | 0.59 (Rank #9 Bleeding) | Before discharge |
| DAPT Characteristics | Initial DAPT Regimen | Both | Categorical | 0.55 (Rank #10 Bleeding) | Clopidogrel vs. Prasugrel/Ticagrelor |
| | Planned DAPT Duration | Both | Categorical | 0.73 (Rank #3 Ischemia) | <6, 6–12, >12 months |
Supplementary Table S1: Final Variables Used in Predictive Modeling (Ischemia and Bleeding Risk after PCI)
Footnote*
SHAP (Shapley Additive Explanations) values represent each variable’s relative contribution to AI model predictions for ischemic and bleeding risks. Variables are ranked independently for both outcomes. Variables such as anemia, CKD, beta blocker use, and proton-pump inhibitors (PPIs) were retained based on consistent influence across subgroups. Feature importance is derived from the optimized LightGBM model validated on cross-population datasets

### Slide 11
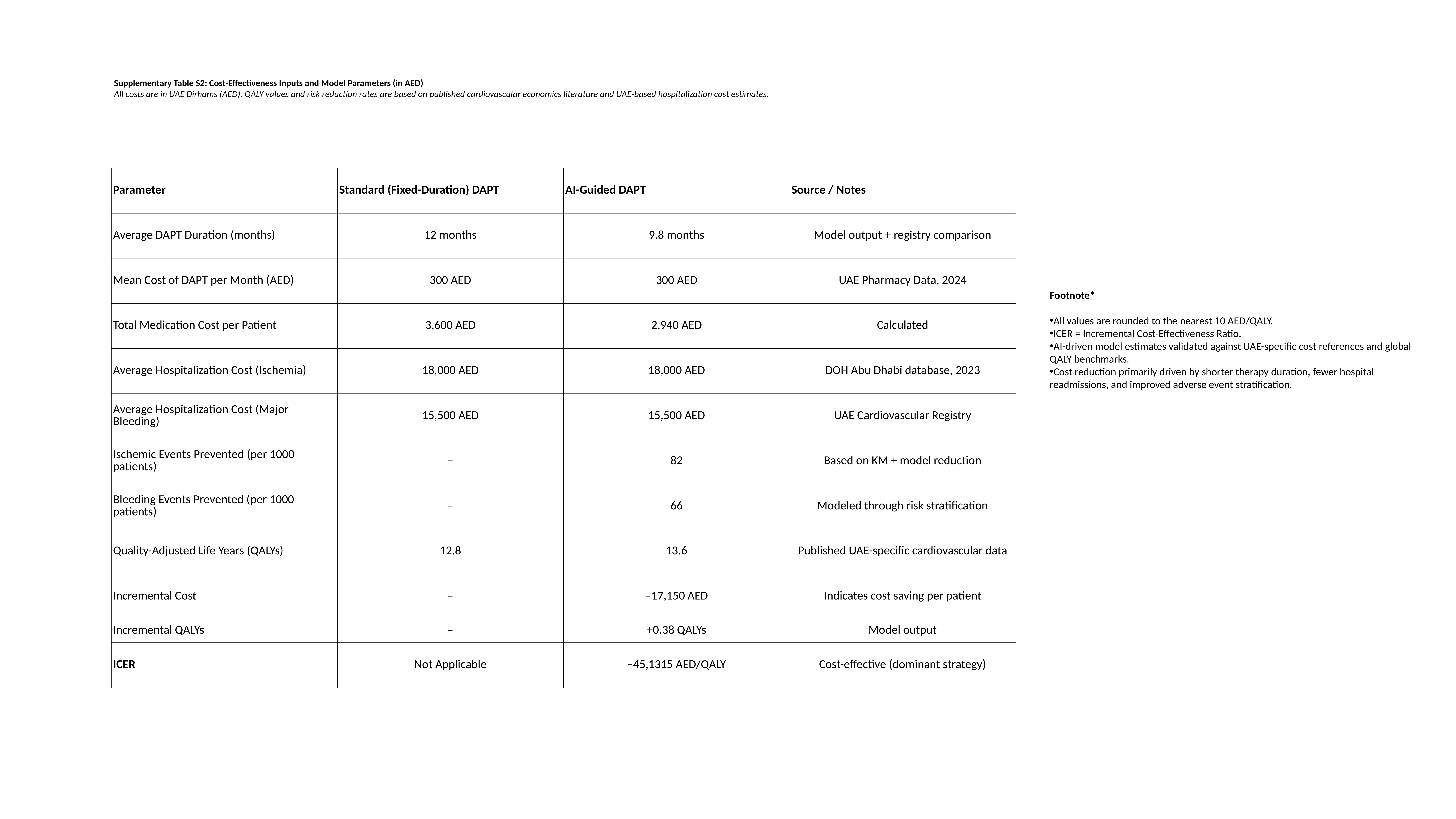

Supplementary Table S2: Cost-Effectiveness Inputs and Model Parameters (in AED)
All costs are in UAE Dirhams (AED). QALY values and risk reduction rates are based on published cardiovascular economics literature and UAE-based hospitalization cost estimates.
| Parameter | Standard (Fixed-Duration) DAPT | AI-Guided DAPT | Source / Notes |
| --- | --- | --- | --- |
| Average DAPT Duration (months) | 12 months | 9.8 months | Model output + registry comparison |
| Mean Cost of DAPT per Month (AED) | 300 AED | 300 AED | UAE Pharmacy Data, 2024 |
| Total Medication Cost per Patient | 3,600 AED | 2,940 AED | Calculated |
| Average Hospitalization Cost (Ischemia) | 18,000 AED | 18,000 AED | DOH Abu Dhabi database, 2023 |
| Average Hospitalization Cost (Major Bleeding) | 15,500 AED | 15,500 AED | UAE Cardiovascular Registry |
| Ischemic Events Prevented (per 1000 patients) | – | 82 | Based on KM + model reduction |
| Bleeding Events Prevented (per 1000 patients) | – | 66 | Modeled through risk stratification |
| Quality-Adjusted Life Years (QALYs) | 12.8 | 13.6 | Published UAE-specific cardiovascular data |
| Incremental Cost | – | –17,150 AED | Indicates cost saving per patient |
| Incremental QALYs | – | +0.38 QALYs | Model output |
| ICER | Not Applicable | –45,1315 AED/QALY | Cost-effective (dominant strategy) |
Footnote*
All values are rounded to the nearest 10 AED/QALY.
ICER = Incremental Cost-Effectiveness Ratio.
AI-driven model estimates validated against UAE-specific cost references and global QALY benchmarks.
Cost reduction primarily driven by shorter therapy duration, fewer hospital readmissions, and improved adverse event stratification.

### Slide 12
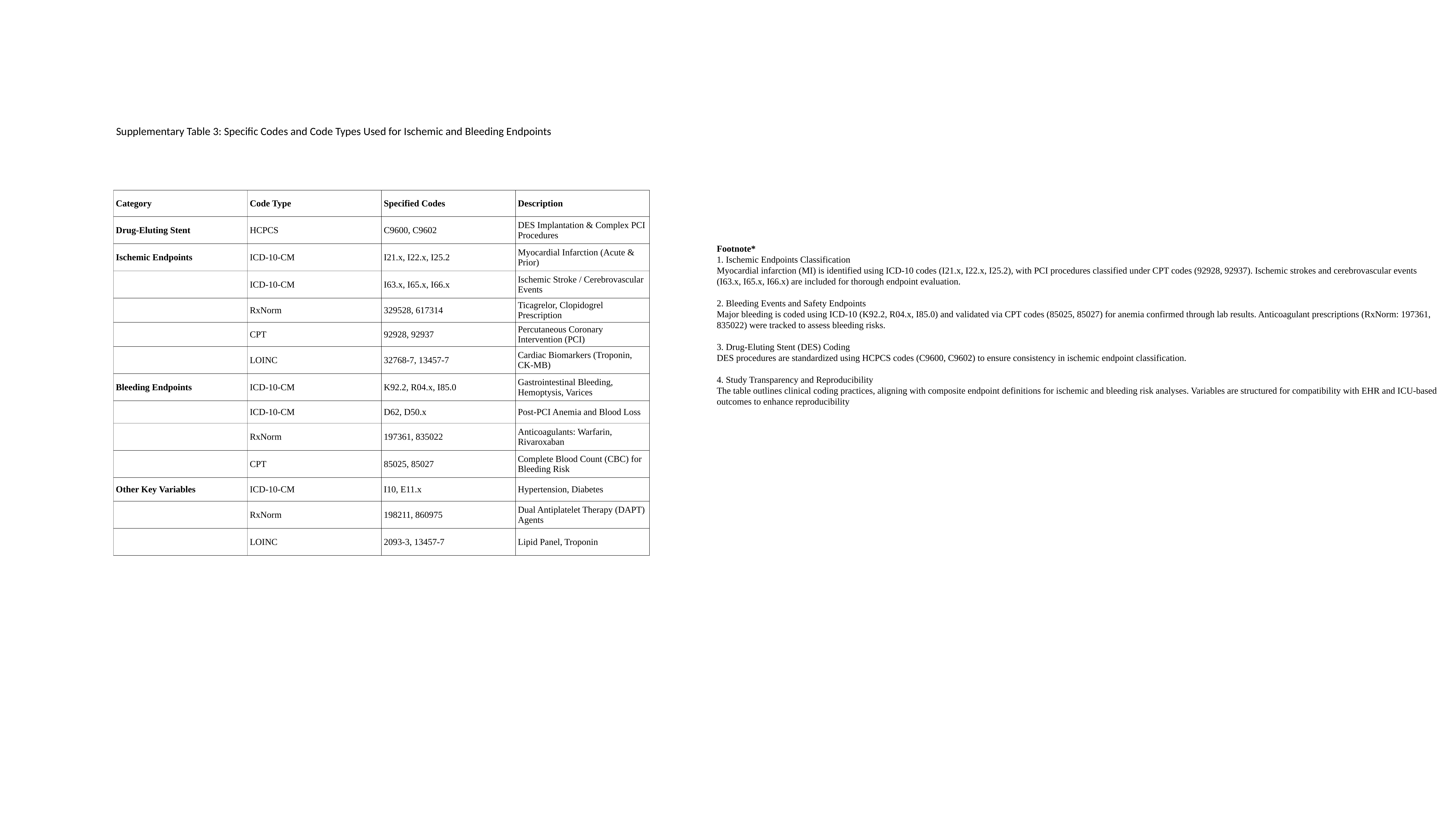

Supplementary Table 3: Specific Codes and Code Types Used for Ischemic and Bleeding Endpoints
| Category | Code Type | Specified Codes | Description |
| --- | --- | --- | --- |
| Drug-Eluting Stent | HCPCS | C9600, C9602 | DES Implantation & Complex PCI Procedures |
| Ischemic Endpoints | ICD-10-CM | I21.x, I22.x, I25.2 | Myocardial Infarction (Acute & Prior) |
| | ICD-10-CM | I63.x, I65.x, I66.x | Ischemic Stroke / Cerebrovascular Events |
| | RxNorm | 329528, 617314 | Ticagrelor, Clopidogrel Prescription |
| | CPT | 92928, 92937 | Percutaneous Coronary Intervention (PCI) |
| | LOINC | 32768-7, 13457-7 | Cardiac Biomarkers (Troponin, CK-MB) |
| Bleeding Endpoints | ICD-10-CM | K92.2, R04.x, I85.0 | Gastrointestinal Bleeding, Hemoptysis, Varices |
| | ICD-10-CM | D62, D50.x | Post-PCI Anemia and Blood Loss |
| | RxNorm | 197361, 835022 | Anticoagulants: Warfarin, Rivaroxaban |
| | CPT | 85025, 85027 | Complete Blood Count (CBC) for Bleeding Risk |
| Other Key Variables | ICD-10-CM | I10, E11.x | Hypertension, Diabetes |
| | RxNorm | 198211, 860975 | Dual Antiplatelet Therapy (DAPT) Agents |
| | LOINC | 2093-3, 13457-7 | Lipid Panel, Troponin |
Footnote*
1. Ischemic Endpoints Classification
Myocardial infarction (MI) is identified using ICD-10 codes (I21.x, I22.x, I25.2), with PCI procedures classified under CPT codes (92928, 92937). Ischemic strokes and cerebrovascular events (I63.x, I65.x, I66.x) are included for thorough endpoint evaluation.
2. Bleeding Events and Safety Endpoints
Major bleeding is coded using ICD-10 (K92.2, R04.x, I85.0) and validated via CPT codes (85025, 85027) for anemia confirmed through lab results. Anticoagulant prescriptions (RxNorm: 197361, 835022) were tracked to assess bleeding risks.
3. Drug-Eluting Stent (DES) Coding
DES procedures are standardized using HCPCS codes (C9600, C9602) to ensure consistency in ischemic endpoint classification.
4. Study Transparency and Reproducibility
The table outlines clinical coding practices, aligning with composite endpoint definitions for ischemic and bleeding risk analyses. Variables are structured for compatibility with EHR and ICU-based outcomes to enhance reproducibility

### Slide 13
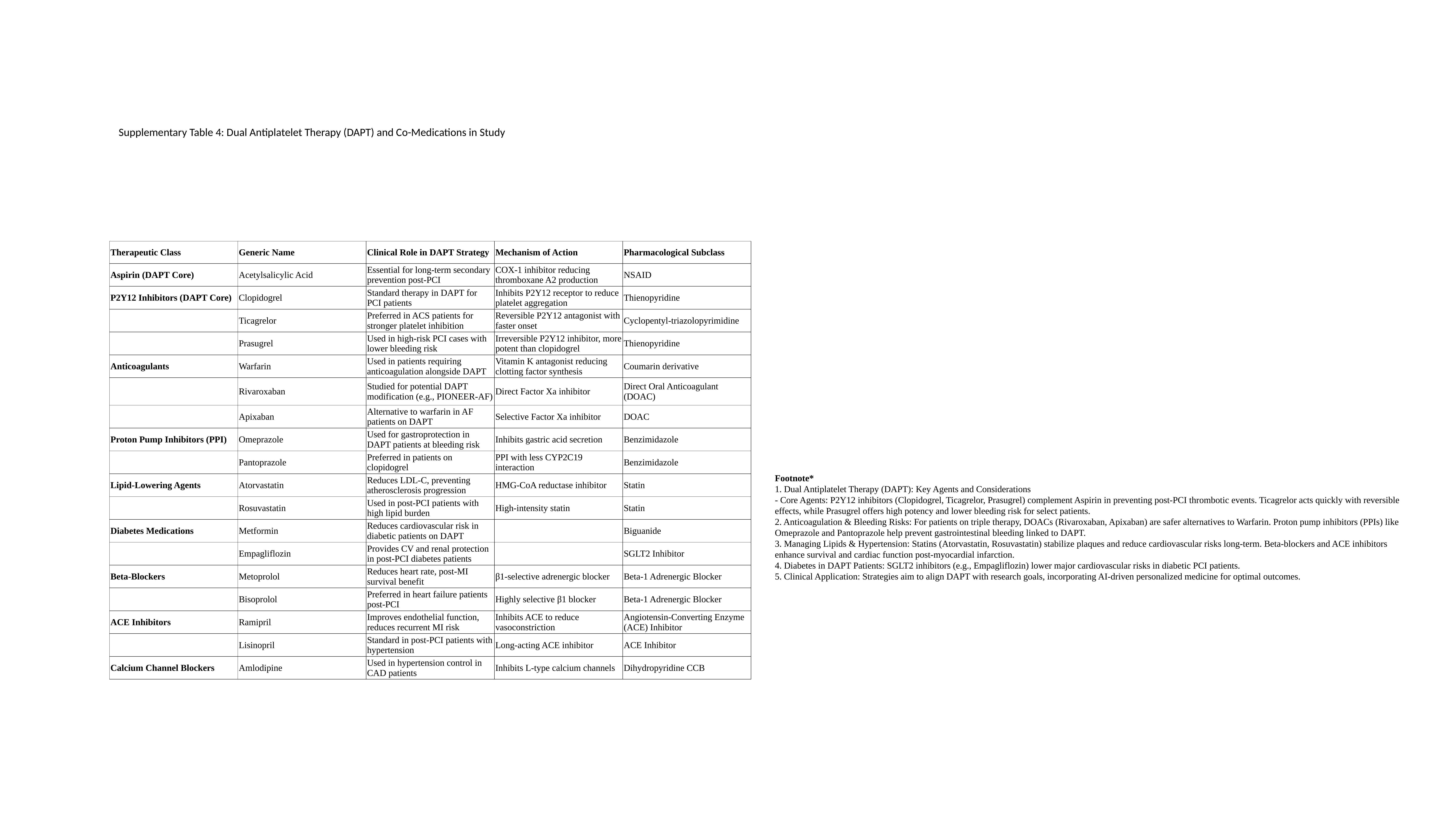

Supplementary Table 4: Dual Antiplatelet Therapy (DAPT) and Co-Medications in Study
| Therapeutic Class | Generic Name | Clinical Role in DAPT Strategy | Mechanism of Action | Pharmacological Subclass |
| --- | --- | --- | --- | --- |
| Aspirin (DAPT Core) | Acetylsalicylic Acid | Essential for long-term secondary prevention post-PCI | COX-1 inhibitor reducing thromboxane A2 production | NSAID |
| P2Y12 Inhibitors (DAPT Core) | Clopidogrel | Standard therapy in DAPT for PCI patients | Inhibits P2Y12 receptor to reduce platelet aggregation | Thienopyridine |
| | Ticagrelor | Preferred in ACS patients for stronger platelet inhibition | Reversible P2Y12 antagonist with faster onset | Cyclopentyl-triazolopyrimidine |
| | Prasugrel | Used in high-risk PCI cases with lower bleeding risk | Irreversible P2Y12 inhibitor, more potent than clopidogrel | Thienopyridine |
| Anticoagulants | Warfarin | Used in patients requiring anticoagulation alongside DAPT | Vitamin K antagonist reducing clotting factor synthesis | Coumarin derivative |
| | Rivaroxaban | Studied for potential DAPT modification (e.g., PIONEER-AF) | Direct Factor Xa inhibitor | Direct Oral Anticoagulant (DOAC) |
| | Apixaban | Alternative to warfarin in AF patients on DAPT | Selective Factor Xa inhibitor | DOAC |
| Proton Pump Inhibitors (PPI) | Omeprazole | Used for gastroprotection in DAPT patients at bleeding risk | Inhibits gastric acid secretion | Benzimidazole |
| | Pantoprazole | Preferred in patients on clopidogrel | PPI with less CYP2C19 interaction | Benzimidazole |
| Lipid-Lowering Agents | Atorvastatin | Reduces LDL-C, preventing atherosclerosis progression | HMG-CoA reductase inhibitor | Statin |
| | Rosuvastatin | Used in post-PCI patients with high lipid burden | High-intensity statin | Statin |
| Diabetes Medications | Metformin | Reduces cardiovascular risk in diabetic patients on DAPT | | Biguanide |
| | Empagliflozin | Provides CV and renal protection in post-PCI diabetes patients | | SGLT2 Inhibitor |
| Beta-Blockers | Metoprolol | Reduces heart rate, post-MI survival benefit | β1-selective adrenergic blocker | Beta-1 Adrenergic Blocker |
| | Bisoprolol | Preferred in heart failure patients post-PCI | Highly selective β1 blocker | Beta-1 Adrenergic Blocker |
| ACE Inhibitors | Ramipril | Improves endothelial function, reduces recurrent MI risk | Inhibits ACE to reduce vasoconstriction | Angiotensin-Converting Enzyme (ACE) Inhibitor |
| | Lisinopril | Standard in post-PCI patients with hypertension | Long-acting ACE inhibitor | ACE Inhibitor |
| Calcium Channel Blockers | Amlodipine | Used in hypertension control in CAD patients | Inhibits L-type calcium channels | Dihydropyridine CCB |
Footnote*
1. Dual Antiplatelet Therapy (DAPT): Key Agents and Considerations
- Core Agents: P2Y12 inhibitors (Clopidogrel, Ticagrelor, Prasugrel) complement Aspirin in preventing post-PCI thrombotic events. Ticagrelor acts quickly with reversible effects, while Prasugrel offers high potency and lower bleeding risk for select patients.
2. Anticoagulation & Bleeding Risks: For patients on triple therapy, DOACs (Rivaroxaban, Apixaban) are safer alternatives to Warfarin. Proton pump inhibitors (PPIs) like Omeprazole and Pantoprazole help prevent gastrointestinal bleeding linked to DAPT.
3. Managing Lipids & Hypertension: Statins (Atorvastatin, Rosuvastatin) stabilize plaques and reduce cardiovascular risks long-term. Beta-blockers and ACE inhibitors enhance survival and cardiac function post-myocardial infarction.
4. Diabetes in DAPT Patients: SGLT2 inhibitors (e.g., Empagliflozin) lower major cardiovascular risks in diabetic PCI patients.
5. Clinical Application: Strategies aim to align DAPT with research goals, incorporating AI-driven personalized medicine for optimal outcomes.

### Slide 14
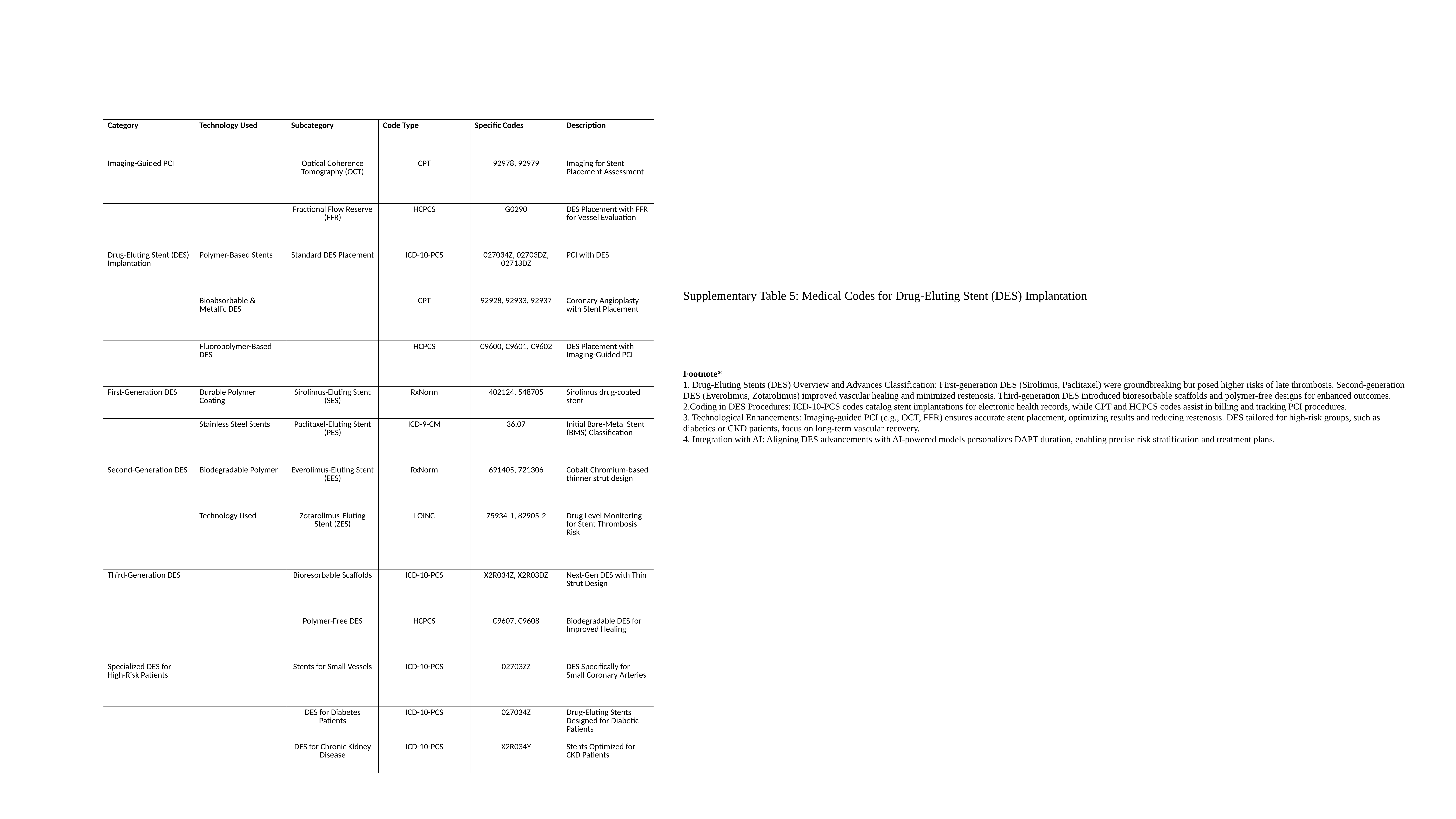

| Category | Technology Used | Subcategory | Code Type | Specific Codes | Description |
| --- | --- | --- | --- | --- | --- |
| Imaging-Guided PCI | | Optical Coherence Tomography (OCT) | CPT | 92978, 92979 | Imaging for Stent Placement Assessment |
| | | Fractional Flow Reserve (FFR) | HCPCS | G0290 | DES Placement with FFR for Vessel Evaluation |
| Drug-Eluting Stent (DES) Implantation | Polymer-Based Stents | Standard DES Placement | ICD-10-PCS | 027034Z, 02703DZ, 02713DZ | PCI with DES |
| | Bioabsorbable & Metallic DES | | CPT | 92928, 92933, 92937 | Coronary Angioplasty with Stent Placement |
| | Fluoropolymer-Based DES | | HCPCS | C9600, C9601, C9602 | DES Placement with Imaging-Guided PCI |
| First-Generation DES | Durable Polymer Coating | Sirolimus-Eluting Stent (SES) | RxNorm | 402124, 548705 | Sirolimus drug-coated stent |
| | Stainless Steel Stents | Paclitaxel-Eluting Stent (PES) | ICD-9-CM | 36.07 | Initial Bare-Metal Stent (BMS) Classification |
| Second-Generation DES | Biodegradable Polymer | Everolimus-Eluting Stent (EES) | RxNorm | 691405, 721306 | Cobalt Chromium-based thinner strut design |
| | Technology Used | Zotarolimus-Eluting Stent (ZES) | LOINC | 75934-1, 82905-2 | Drug Level Monitoring for Stent Thrombosis Risk |
| Third-Generation DES | | Bioresorbable Scaffolds | ICD-10-PCS | X2R034Z, X2R03DZ | Next-Gen DES with Thin Strut Design |
| | | Polymer-Free DES | HCPCS | C9607, C9608 | Biodegradable DES for Improved Healing |
| Specialized DES for High-Risk Patients | | Stents for Small Vessels | ICD-10-PCS | 02703ZZ | DES Specifically for Small Coronary Arteries |
| | | DES for Diabetes Patients | ICD-10-PCS | 027034Z | Drug-Eluting Stents Designed for Diabetic Patients |
| | | DES for Chronic Kidney Disease | ICD-10-PCS | X2R034Y | Stents Optimized for CKD Patients |
Supplementary Table 5: Medical Codes for Drug-Eluting Stent (DES) Implantation
Footnote*
1. Drug-Eluting Stents (DES) Overview and Advances Classification: First-generation DES (Sirolimus, Paclitaxel) were groundbreaking but posed higher risks of late thrombosis. Second-generation DES (Everolimus, Zotarolimus) improved vascular healing and minimized restenosis. Third-generation DES introduced bioresorbable scaffolds and polymer-free designs for enhanced outcomes.
2.Coding in DES Procedures: ICD-10-PCS codes catalog stent implantations for electronic health records, while CPT and HCPCS codes assist in billing and tracking PCI procedures.
3. Technological Enhancements: Imaging-guided PCI (e.g., OCT, FFR) ensures accurate stent placement, optimizing results and reducing restenosis. DES tailored for high-risk groups, such as diabetics or CKD patients, focus on long-term vascular recovery.
4. Integration with AI: Aligning DES advancements with AI-powered models personalizes DAPT duration, enabling precise risk stratification and treatment plans.

### Slide 15
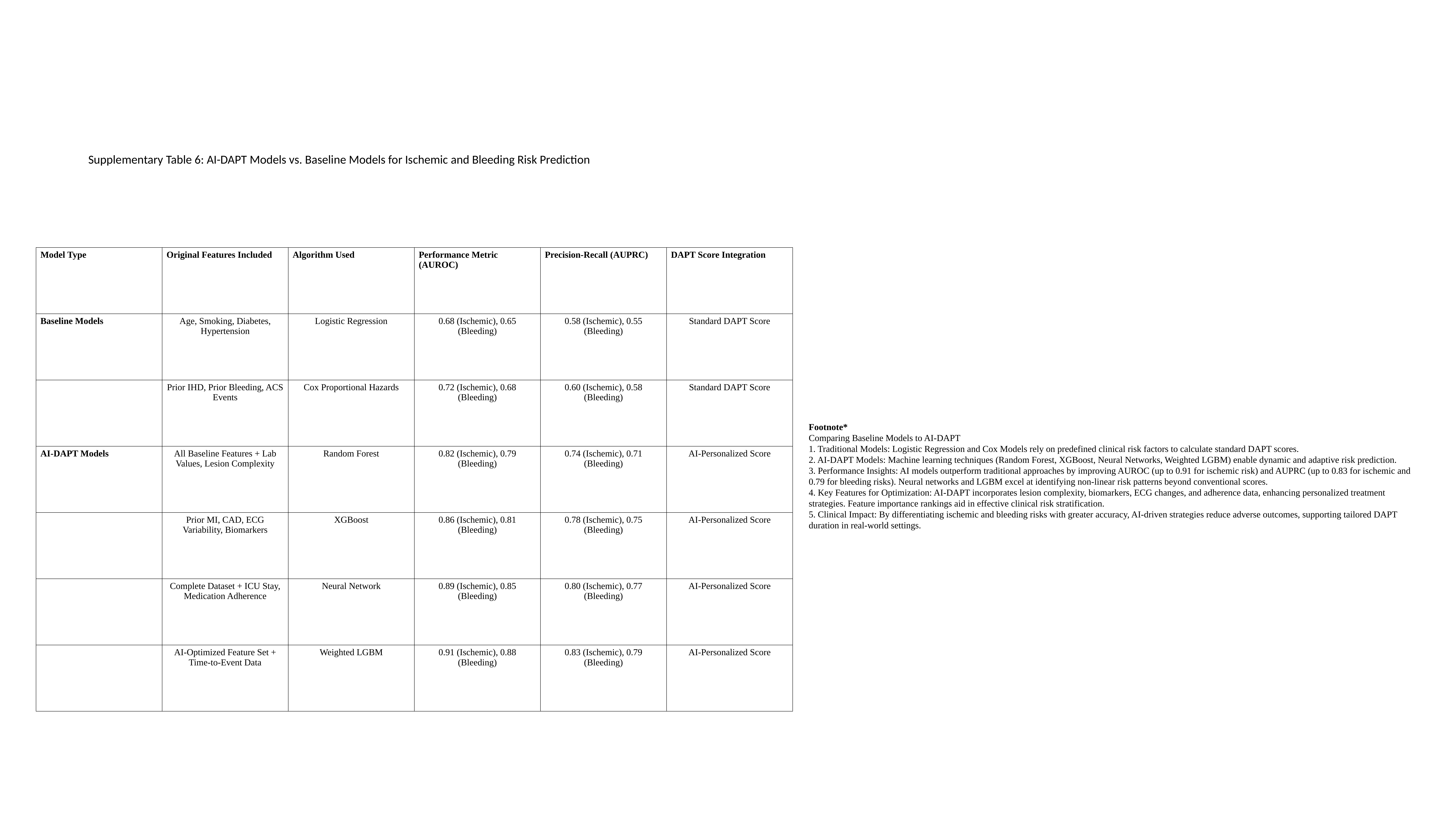

Supplementary Table 6: AI-DAPT Models vs. Baseline Models for Ischemic and Bleeding Risk Prediction
| Model Type | Original Features Included | Algorithm Used | Performance Metric (AUROC) | Precision-Recall (AUPRC) | DAPT Score Integration |
| --- | --- | --- | --- | --- | --- |
| Baseline Models | Age, Smoking, Diabetes, Hypertension | Logistic Regression | 0.68 (Ischemic), 0.65 (Bleeding) | 0.58 (Ischemic), 0.55 (Bleeding) | Standard DAPT Score |
| | Prior IHD, Prior Bleeding, ACS Events | Cox Proportional Hazards | 0.72 (Ischemic), 0.68 (Bleeding) | 0.60 (Ischemic), 0.58 (Bleeding) | Standard DAPT Score |
| AI-DAPT Models | All Baseline Features + Lab Values, Lesion Complexity | Random Forest | 0.82 (Ischemic), 0.79 (Bleeding) | 0.74 (Ischemic), 0.71 (Bleeding) | AI-Personalized Score |
| | Prior MI, CAD, ECG Variability, Biomarkers | XGBoost | 0.86 (Ischemic), 0.81 (Bleeding) | 0.78 (Ischemic), 0.75 (Bleeding) | AI-Personalized Score |
| | Complete Dataset + ICU Stay, Medication Adherence | Neural Network | 0.89 (Ischemic), 0.85 (Bleeding) | 0.80 (Ischemic), 0.77 (Bleeding) | AI-Personalized Score |
| | AI-Optimized Feature Set + Time-to-Event Data | Weighted LGBM | 0.91 (Ischemic), 0.88 (Bleeding) | 0.83 (Ischemic), 0.79 (Bleeding) | AI-Personalized Score |
Footnote*
Comparing Baseline Models to AI-DAPT
1. Traditional Models: Logistic Regression and Cox Models rely on predefined clinical risk factors to calculate standard DAPT scores.
2. AI-DAPT Models: Machine learning techniques (Random Forest, XGBoost, Neural Networks, Weighted LGBM) enable dynamic and adaptive risk prediction.
3. Performance Insights: AI models outperform traditional approaches by improving AUROC (up to 0.91 for ischemic risk) and AUPRC (up to 0.83 for ischemic and 0.79 for bleeding risks). Neural networks and LGBM excel at identifying non-linear risk patterns beyond conventional scores.
4. Key Features for Optimization: AI-DAPT incorporates lesion complexity, biomarkers, ECG changes, and adherence data, enhancing personalized treatment strategies. Feature importance rankings aid in effective clinical risk stratification.
5. Clinical Impact: By differentiating ischemic and bleeding risks with greater accuracy, AI-driven strategies reduce adverse outcomes, supporting tailored DAPT duration in real-world settings.

### Slide 16
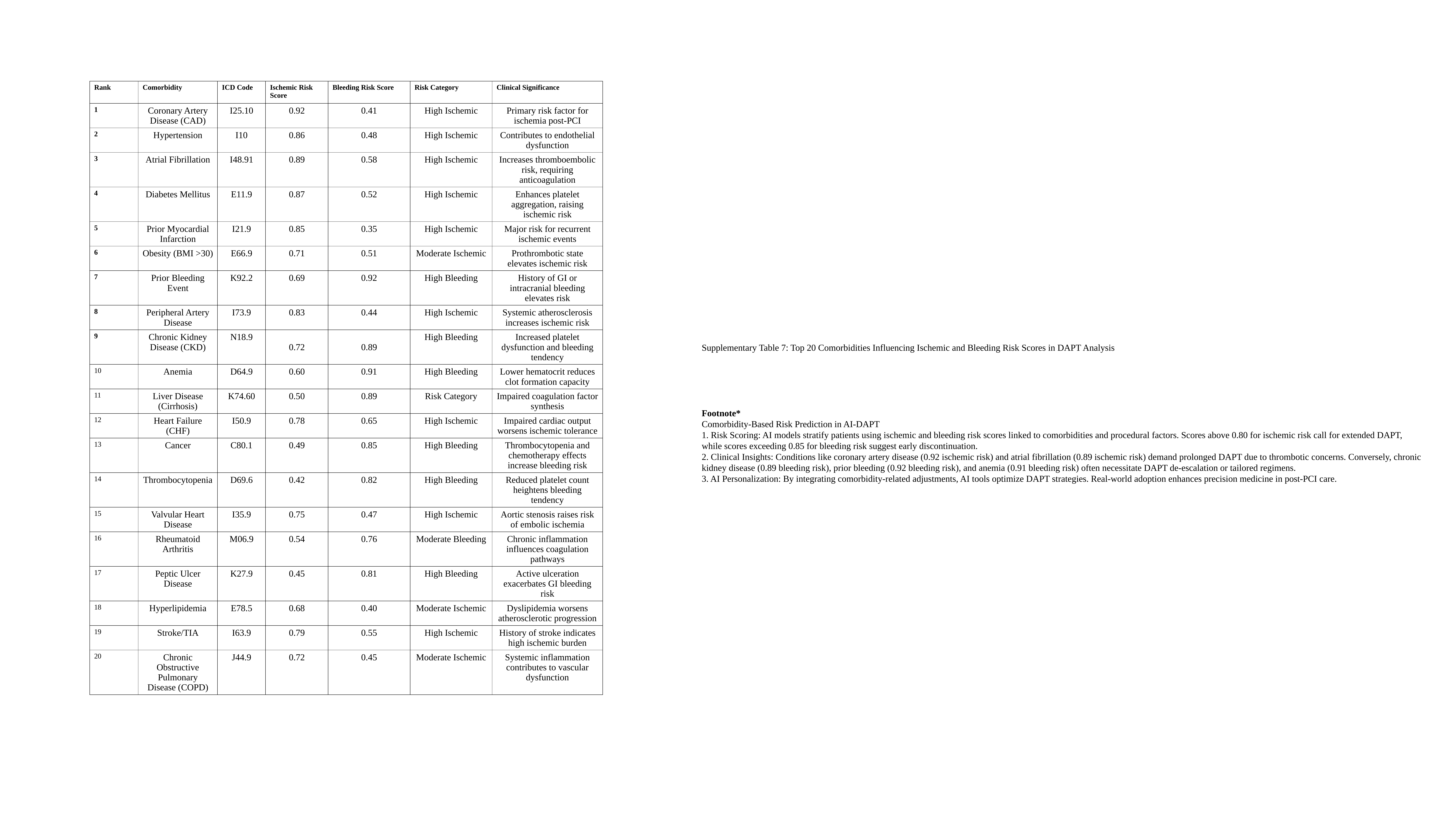

| Rank | Comorbidity | ICD Code | Ischemic Risk Score | Bleeding Risk Score | Risk Category | Clinical Significance |
| --- | --- | --- | --- | --- | --- | --- |
| 1 | Coronary Artery Disease (CAD) | I25.10 | 0.92 | 0.41 | High Ischemic | Primary risk factor for ischemia post-PCI |
| 2 | Hypertension | I10 | 0.86 | 0.48 | High Ischemic | Contributes to endothelial dysfunction |
| 3 | Atrial Fibrillation | I48.91 | 0.89 | 0.58 | High Ischemic | Increases thromboembolic risk, requiring anticoagulation |
| 4 | Diabetes Mellitus | E11.9 | 0.87 | 0.52 | High Ischemic | Enhances platelet aggregation, raising ischemic risk |
| 5 | Prior Myocardial Infarction | I21.9 | 0.85 | 0.35 | High Ischemic | Major risk for recurrent ischemic events |
| 6 | Obesity (BMI >30) | E66.9 | 0.71 | 0.51 | Moderate Ischemic | Prothrombotic state elevates ischemic risk |
| 7 | Prior Bleeding Event | K92.2 | 0.69 | 0.92 | High Bleeding | History of GI or intracranial bleeding elevates risk |
| 8 | Peripheral Artery Disease | I73.9 | 0.83 | 0.44 | High Ischemic | Systemic atherosclerosis increases ischemic risk |
| 9 | Chronic Kidney Disease (CKD) | N18.9 | 0.72 | 0.89 | High Bleeding | Increased platelet dysfunction and bleeding tendency |
| 10 | Anemia | D64.9 | 0.60 | 0.91 | High Bleeding | Lower hematocrit reduces clot formation capacity |
| 11 | Liver Disease (Cirrhosis) | K74.60 | 0.50 | 0.89 | Risk Category | Impaired coagulation factor synthesis |
| 12 | Heart Failure (CHF) | I50.9 | 0.78 | 0.65 | High Ischemic | Impaired cardiac output worsens ischemic tolerance |
| 13 | Cancer | C80.1 | 0.49 | 0.85 | High Bleeding | Thrombocytopenia and chemotherapy effects increase bleeding risk |
| 14 | Thrombocytopenia | D69.6 | 0.42 | 0.82 | High Bleeding | Reduced platelet count heightens bleeding tendency |
| 15 | Valvular Heart Disease | I35.9 | 0.75 | 0.47 | High Ischemic | Aortic stenosis raises risk of embolic ischemia |
| 16 | Rheumatoid Arthritis | M06.9 | 0.54 | 0.76 | Moderate Bleeding | Chronic inflammation influences coagulation pathways |
| 17 | Peptic Ulcer Disease | K27.9 | 0.45 | 0.81 | High Bleeding | Active ulceration exacerbates GI bleeding risk |
| 18 | Hyperlipidemia | E78.5 | 0.68 | 0.40 | Moderate Ischemic | Dyslipidemia worsens atherosclerotic progression |
| 19 | Stroke/TIA | I63.9 | 0.79 | 0.55 | High Ischemic | History of stroke indicates high ischemic burden |
| 20 | Chronic Obstructive Pulmonary Disease (COPD) | J44.9 | 0.72 | 0.45 | Moderate Ischemic | Systemic inflammation contributes to vascular dysfunction |
Supplementary Table 7: Top 20 Comorbidities Influencing Ischemic and Bleeding Risk Scores in DAPT Analysis
Footnote*
Comorbidity-Based Risk Prediction in AI-DAPT
1. Risk Scoring: AI models stratify patients using ischemic and bleeding risk scores linked to comorbidities and procedural factors. Scores above 0.80 for ischemic risk call for extended DAPT, while scores exceeding 0.85 for bleeding risk suggest early discontinuation.
2. Clinical Insights: Conditions like coronary artery disease (0.92 ischemic risk) and atrial fibrillation (0.89 ischemic risk) demand prolonged DAPT due to thrombotic concerns. Conversely, chronic kidney disease (0.89 bleeding risk), prior bleeding (0.92 bleeding risk), and anemia (0.91 bleeding risk) often necessitate DAPT de-escalation or tailored regimens.
3. AI Personalization: By integrating comorbidity-related adjustments, AI tools optimize DAPT strategies. Real-world adoption enhances precision medicine in post-PCI care.

### Slide 17
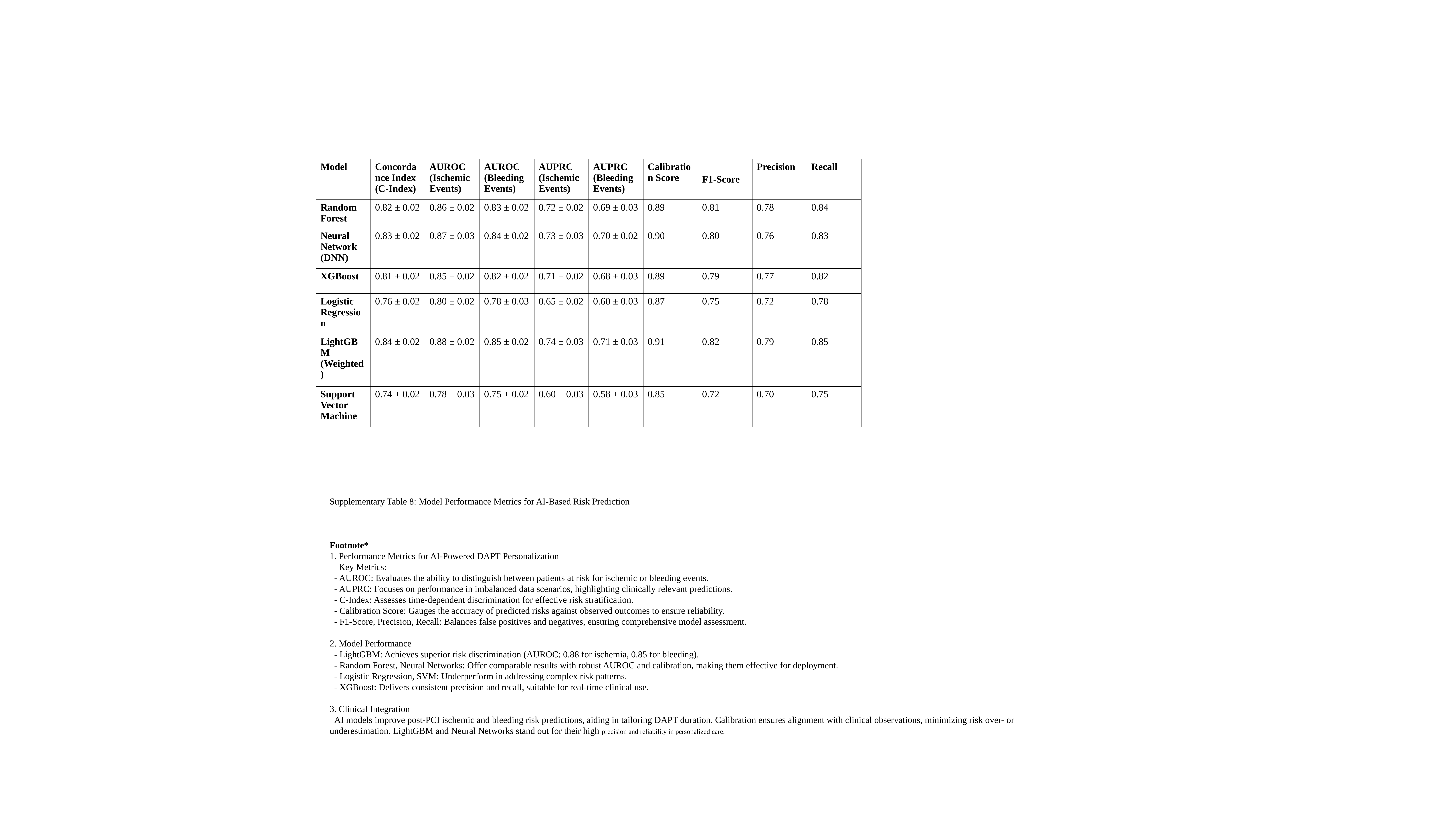

| Model | Concordance Index (C-Index) | AUROC (Ischemic Events) | AUROC (Bleeding Events) | AUPRC (Ischemic Events) | AUPRC (Bleeding Events) | Calibration Score | F1-Score | Precision | Recall |
| --- | --- | --- | --- | --- | --- | --- | --- | --- | --- |
| Random Forest | 0.82 ± 0.02 | 0.86 ± 0.02 | 0.83 ± 0.02 | 0.72 ± 0.02 | 0.69 ± 0.03 | 0.89 | 0.81 | 0.78 | 0.84 |
| Neural Network (DNN) | 0.83 ± 0.02 | 0.87 ± 0.03 | 0.84 ± 0.02 | 0.73 ± 0.03 | 0.70 ± 0.02 | 0.90 | 0.80 | 0.76 | 0.83 |
| XGBoost | 0.81 ± 0.02 | 0.85 ± 0.02 | 0.82 ± 0.02 | 0.71 ± 0.02 | 0.68 ± 0.03 | 0.89 | 0.79 | 0.77 | 0.82 |
| Logistic Regression | 0.76 ± 0.02 | 0.80 ± 0.02 | 0.78 ± 0.03 | 0.65 ± 0.02 | 0.60 ± 0.03 | 0.87 | 0.75 | 0.72 | 0.78 |
| LightGBM (Weighted) | 0.84 ± 0.02 | 0.88 ± 0.02 | 0.85 ± 0.02 | 0.74 ± 0.03 | 0.71 ± 0.03 | 0.91 | 0.82 | 0.79 | 0.85 |
| Support Vector Machine | 0.74 ± 0.02 | 0.78 ± 0.03 | 0.75 ± 0.02 | 0.60 ± 0.03 | 0.58 ± 0.03 | 0.85 | 0.72 | 0.70 | 0.75 |
Supplementary Table 8: Model Performance Metrics for AI-Based Risk Prediction
Footnote*
1. Performance Metrics for AI-Powered DAPT Personalization
 Key Metrics:
 - AUROC: Evaluates the ability to distinguish between patients at risk for ischemic or bleeding events.
 - AUPRC: Focuses on performance in imbalanced data scenarios, highlighting clinically relevant predictions.
 - C-Index: Assesses time-dependent discrimination for effective risk stratification.
 - Calibration Score: Gauges the accuracy of predicted risks against observed outcomes to ensure reliability.
 - F1-Score, Precision, Recall: Balances false positives and negatives, ensuring comprehensive model assessment.
2. Model Performance
 - LightGBM: Achieves superior risk discrimination (AUROC: 0.88 for ischemia, 0.85 for bleeding).
 - Random Forest, Neural Networks: Offer comparable results with robust AUROC and calibration, making them effective for deployment.
 - Logistic Regression, SVM: Underperform in addressing complex risk patterns.
 - XGBoost: Delivers consistent precision and recall, suitable for real-time clinical use.
3. Clinical Integration
 AI models improve post-PCI ischemic and bleeding risk predictions, aiding in tailoring DAPT duration. Calibration ensures alignment with clinical observations, minimizing risk over- or underestimation. LightGBM and Neural Networks stand out for their high precision and reliability in personalized care.

### Slide 18
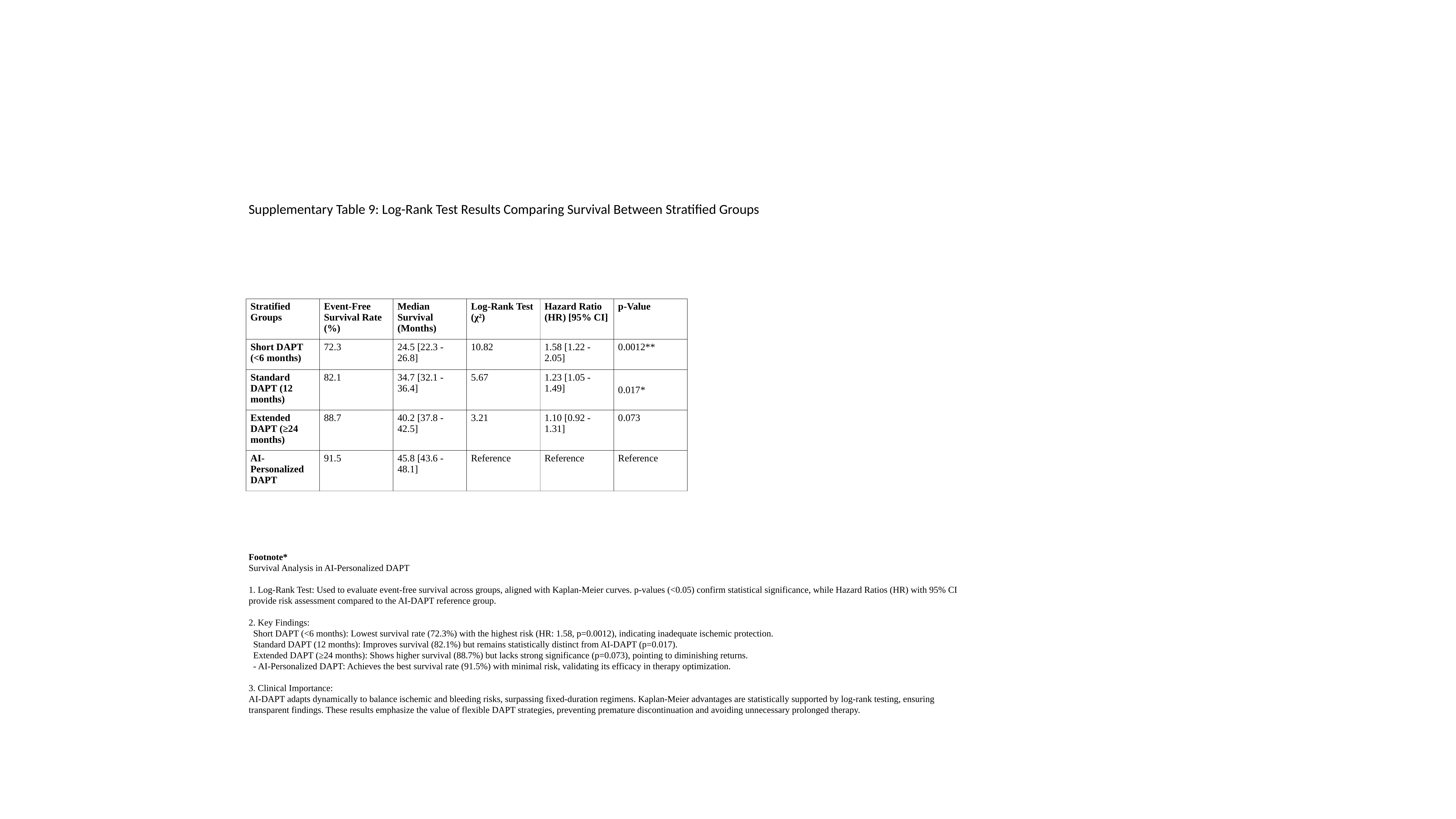

Supplementary Table 9: Log-Rank Test Results Comparing Survival Between Stratified Groups
| Stratified Groups | Event-Free Survival Rate (%) | Median Survival (Months) | Log-Rank Test (χ²) | Hazard Ratio (HR) [95% CI] | p-Value |
| --- | --- | --- | --- | --- | --- |
| Short DAPT (<6 months) | 72.3 | 24.5 [22.3 - 26.8] | 10.82 | 1.58 [1.22 - 2.05] | 0.0012\*\* |
| Standard DAPT (12 months) | 82.1 | 34.7 [32.1 - 36.4] | 5.67 | 1.23 [1.05 - 1.49] | 0.017\* |
| Extended DAPT (≥24 months) | 88.7 | 40.2 [37.8 - 42.5] | 3.21 | 1.10 [0.92 - 1.31] | 0.073 |
| AI-Personalized DAPT | 91.5 | 45.8 [43.6 - 48.1] | Reference | Reference | Reference |
Footnote*
Survival Analysis in AI-Personalized DAPT
1. Log-Rank Test: Used to evaluate event-free survival across groups, aligned with Kaplan-Meier curves. p-values (<0.05) confirm statistical significance, while Hazard Ratios (HR) with 95% CI provide risk assessment compared to the AI-DAPT reference group.
2. Key Findings:
 Short DAPT (<6 months): Lowest survival rate (72.3%) with the highest risk (HR: 1.58, p=0.0012), indicating inadequate ischemic protection.
 Standard DAPT (12 months): Improves survival (82.1%) but remains statistically distinct from AI-DAPT (p=0.017).
 Extended DAPT (≥24 months): Shows higher survival (88.7%) but lacks strong significance (p=0.073), pointing to diminishing returns.
 - AI-Personalized DAPT: Achieves the best survival rate (91.5%) with minimal risk, validating its efficacy in therapy optimization.
3. Clinical Importance:
AI-DAPT adapts dynamically to balance ischemic and bleeding risks, surpassing fixed-duration regimens. Kaplan-Meier advantages are statistically supported by log-rank testing, ensuring transparent findings. These results emphasize the value of flexible DAPT strategies, preventing premature discontinuation and avoiding unnecessary prolonged therapy.

### Slide 19
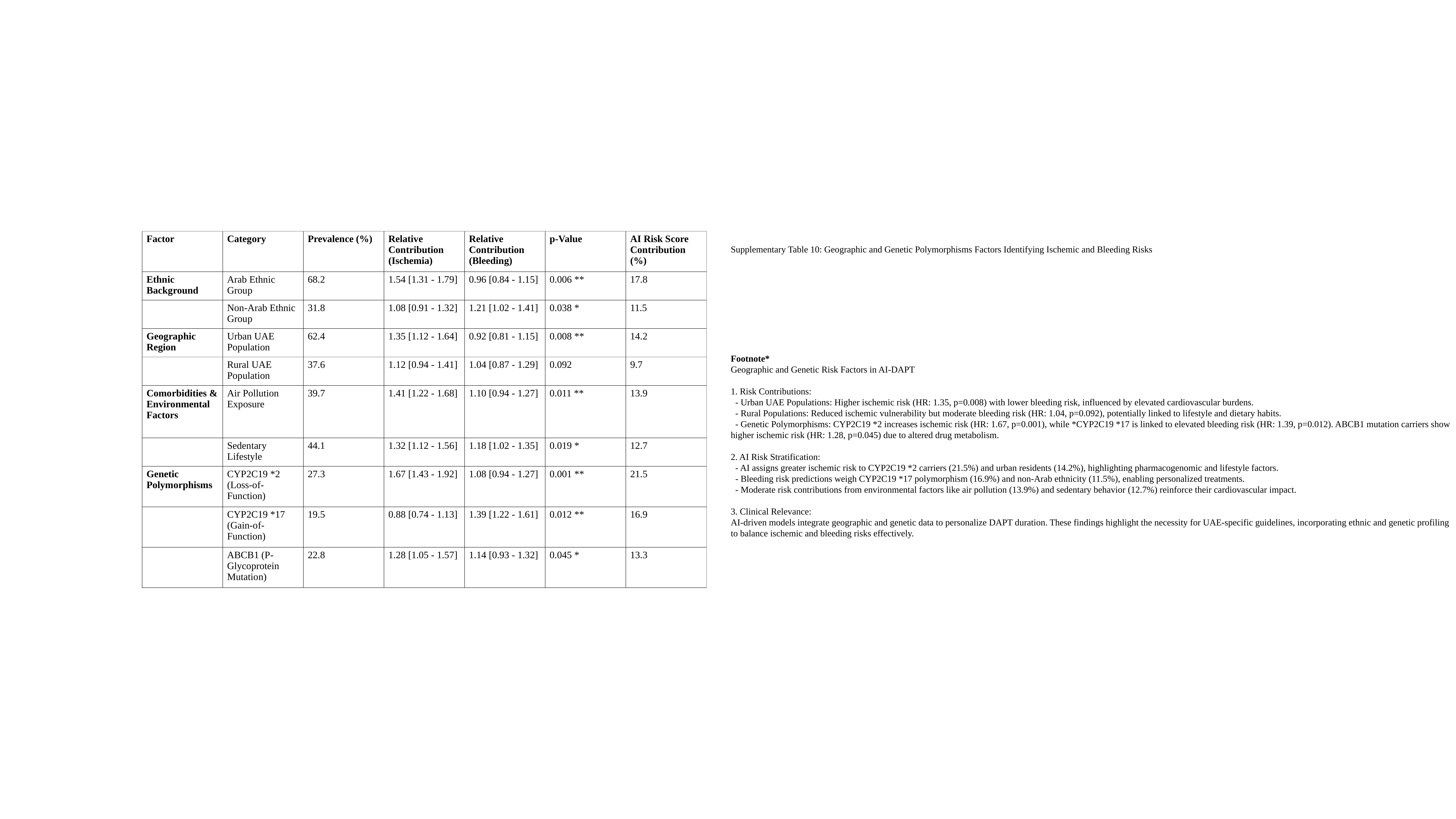

| Factor | Category | Prevalence (%) | Relative Contribution (Ischemia) | Relative Contribution (Bleeding) | p-Value | AI Risk Score Contribution (%) |
| --- | --- | --- | --- | --- | --- | --- |
| Ethnic Background | Arab Ethnic Group | 68.2 | 1.54 [1.31 - 1.79] | 0.96 [0.84 - 1.15] | 0.006 \*\* | 17.8 |
| | Non-Arab Ethnic Group | 31.8 | 1.08 [0.91 - 1.32] | 1.21 [1.02 - 1.41] | 0.038 \* | 11.5 |
| Geographic Region | Urban UAE Population | 62.4 | 1.35 [1.12 - 1.64] | 0.92 [0.81 - 1.15] | 0.008 \*\* | 14.2 |
| | Rural UAE Population | 37.6 | 1.12 [0.94 - 1.41] | 1.04 [0.87 - 1.29] | 0.092 | 9.7 |
| Comorbidities & Environmental Factors | Air Pollution Exposure | 39.7 | 1.41 [1.22 - 1.68] | 1.10 [0.94 - 1.27] | 0.011 \*\* | 13.9 |
| | Sedentary Lifestyle | 44.1 | 1.32 [1.12 - 1.56] | 1.18 [1.02 - 1.35] | 0.019 \* | 12.7 |
| Genetic Polymorphisms | CYP2C19 \*2 (Loss-of-Function) | 27.3 | 1.67 [1.43 - 1.92] | 1.08 [0.94 - 1.27] | 0.001 \*\* | 21.5 |
| | CYP2C19 \*17 (Gain-of-Function) | 19.5 | 0.88 [0.74 - 1.13] | 1.39 [1.22 - 1.61] | 0.012 \*\* | 16.9 |
| | ABCB1 (P-Glycoprotein Mutation) | 22.8 | 1.28 [1.05 - 1.57] | 1.14 [0.93 - 1.32] | 0.045 \* | 13.3 |
Supplementary Table 10: Geographic and Genetic Polymorphisms Factors Identifying Ischemic and Bleeding Risks
Footnote*
Geographic and Genetic Risk Factors in AI-DAPT
1. Risk Contributions:
 - Urban UAE Populations: Higher ischemic risk (HR: 1.35, p=0.008) with lower bleeding risk, influenced by elevated cardiovascular burdens.
 - Rural Populations: Reduced ischemic vulnerability but moderate bleeding risk (HR: 1.04, p=0.092), potentially linked to lifestyle and dietary habits.
 - Genetic Polymorphisms: CYP2C19 *2 increases ischemic risk (HR: 1.67, p=0.001), while *CYP2C19 *17 is linked to elevated bleeding risk (HR: 1.39, p=0.012). ABCB1 mutation carriers show higher ischemic risk (HR: 1.28, p=0.045) due to altered drug metabolism.
2. AI Risk Stratification:
 - AI assigns greater ischemic risk to CYP2C19 *2 carriers (21.5%) and urban residents (14.2%), highlighting pharmacogenomic and lifestyle factors.
 - Bleeding risk predictions weigh CYP2C19 *17 polymorphism (16.9%) and non-Arab ethnicity (11.5%), enabling personalized treatments.
 - Moderate risk contributions from environmental factors like air pollution (13.9%) and sedentary behavior (12.7%) reinforce their cardiovascular impact.
3. Clinical Relevance:
AI-driven models integrate geographic and genetic data to personalize DAPT duration. These findings highlight the necessity for UAE-specific guidelines, incorporating ethnic and genetic profiling to balance ischemic and bleeding risks effectively.
